# The influence of CYP2D6 and CYP2C19 genetic variation on diabetes mellitus risk in people taking antidepressants and antipsychotics

**DOI:** 10.1101/2021.07.07.21259926

**Authors:** Isabelle Austin-Zimmerman, Marta Wronska, Baihan Wang, Haritz Irizar, Johan Hilge Thygesen, Anjali Bhat, Spiros Denaxas, Ghazaleh Fatemifar, Chris Finan, Jasmine Harju-Seppänen, Olga Giannakopoulou, Karoline Kuchenbaecker, Eirini Zartaloudi, Andrew McQuillin, Elvira Bramon

**Affiliations:** Division of Psychiatry, University College London; Department of Genetics & Genomic Sciences, Icahn School of Medicine at Mount Sinai, New York, NY, USA; Institute of Health Informatics, University College London; HDR UK, Institute of Health Informatics, University College London; Institute of Cardiovascular Science, Faculty of Population Health, University College London; UCL British Heart Foundation Research Accelerator; Department of Cardiology, Division Heart and Lungs, University Medical Center Utrecht, Heidelberglaan 100, 3584 CX Utrecht, the Netherlands; Genetics Institute, University College London

## Abstract

**Background:** CYP2D6 and CYP2C19 enzymes are essential in the metabolism of antidepressants and antipsychotics. Genetic variation in these genes may increase risk of adverse drug reactions. Antidepressants and antipsychotics have previously been associated with risk of diabetes. We examined whether individual genetic differences in CYP2D6 and CYP2C19 contribute to these effects.

**Methods:** We identified 31,579 individuals taking antidepressants and 2,699 taking antipsychotics within UK Biobank. Participants were classified as poor, intermediate or normal metabolisers of CYP2D6, and as poor, intermediate, normal, rapid and ultra-rapid metabolisers of CYP2C19. Risk of diabetes mellitus represented by HbA1c level was examined in relation to the metabolic phenotypes. We analysed drugs either individually (where sample size permitted) or grouped by class.

**Results:** CYP2D6 poor metabolisers taking paroxetine had higher Hb1Ac than normal metabolisers (mean difference: 2.29mmol/mol; p < 0.001). Among participants with diabetes who were taking venlafaxine, CYP2D6 poor metabolisers had higher HbA1c levels compared to normal metabolisers (mean differences: 10.15 mmol/mol; p < 0.001. Among participants with diabetes who were taking fluoxetine, we observe that CYP2D6 intermediate metabolisers and decreased HbA1c, compared to normal metabolisers (mean difference - 7.74mmol/mol; p=0.017). We did not observe any relationship between CYP2D6 or CYP2C19 metabolic status and HbA1c levels in participants taking antipsychotic medication.

**Conclusion:** Our results indicate that the impact of genetic variation in CYP2D6 differs depending on diabetes status. Although our findings support existing clinical guidelines, further research is essential to inform pharmacogenetic testing for people taking antidepressants and antipsychotics.

## Introduction

The use of both antidepressant and antipsychotic medications has increased steadily in recent years. Antidepressant drugs were the third most commonly prescribed drug group in 2018, with 70.9 million prescriptions across the United Kingdom – an almost two-fold increase since 2008 (1,2). It is estimated that almost 20% of the British adult population has been prescribed an antidepressant at some stage (1–3). A similar trend is seen in the prescription of antipsychotics, with an increase from eight to 12 million prescriptions between 2008 and 2018 (2). Both antidepressant and antipsychotic medication provide essential and often lifesaving treatment for many patients. However, they are also associated with a range of common and sometimes serious adverse drug reactions including sedation, weight gain, movement disorders, and an increased risk of developing diabetes mellitus (4,5).

Most first-generation antipsychotics, as well as olanzapine and clozapine, have been shown to impair glucose regulation (5–10). Other second generation (or atypical) antipsychotics such as amisulpride, ziprasidone, and aripiprazole seem less associated with this risk (5–10). Several studies have linked tricyclic antidepressants to increased diabetes risk (4,11–13). The evidence for selective serotonin reuptake inhibitors (SSRIs) is inconsistent, with some studies showing improved diabetic control and others showing the opposite (4,11). Research into serotonin-noradrenaline reuptake inhibitors (SNRIs), such as venlafaxine, has reported both a lack of influence on glycaemic control and diabetes risk (10,14–16). Some research suggests that the risk of antidepressant-induced diabetes varies substantially between similar drugs of the same class, and thus may not be a mechanism-based adverse effect, but rather an off-target effect of a single drug (17).

Pharmacogenetics may help explain inter-individual differences in drug response and adverse drug reactions. Cytochrome P450 (CYP450) is a superfamily of enzymes involved in the oxidative biotransformation and clearance of the majority of prescribed drugs (18). CYP2D6 and CYP2C19 are the two CYP450 enzymes most involved in the metabolism of antidepressant and antipsychotic drugs and are both highly polymorphic (18,19). Genetic variation in these genes results in an altered enzyme activity and thus may explain some of the interindividual differences in treatment response. Typically, individuals are grouped into four to five phenotypic groups reflecting differing metabolic capabilities (19,20). Poor metabolisers lack a functional enzyme due to defective or deleted genes; intermediate metabolisers usually have one functional and one defective or deleted allele causing reduced activity of the enzyme; rapid and ultra-rapid metabolisers usually have multiple copies of a functional gene or possess variants that increase gene expression (21). Normal metabolisers (previously described as ‘extensive metabolisers’), or wild-type, are those with two fully functional copies of the gene and thus ‘normal’ enzymatic activity. The prevalence of CYP2D6 and CYP2C19 phenotypes varies across populations, but the extreme metabolisers are typically the least commonly observed: less than 10% of people are poor metabolisers, and less than 3% are ultra-rapid metabolisers, across all major populations and for both genes (22,23).

Several studies have shown that poor metabolisers of CYP2D6 or CYP2C19 have higher serum levels of antidepressants and antipsychotics, compared to normal metabolisers (24–30). The Clinical Pharmacogenomics Implementation Consortium (CPIC) has developed evidence-based clinical guidelines for SSRIs and tricyclic antidepressants, recommending adjusted dosing based on CYP2D6 and CYP2C19 metabolic status (31,32). There are currently no CPIC guidelines for antipsychotics, but the Dutch Pharmacogenetics Working Group provides guidelines for aripiprazole, haloperidol, pimozide and zuclopenthixol based on CYP2D6 genotype (33). Work to incorporate similar evidence based clinical guidelines to the UK National Health Service (NHS) is ongoing (34).

Thus far, research on the putative association between CYP450 metabolic phenotype and adverse drug reactions in response to antidepressants and antipsychotics has been limited by small sample sizes (34,35). Little is known about pharmacogenetic influences on the diabetes risk associated with these drugs. Therefore, this study aims to examine the association between CYP2C19 and CYP2D6 metabolic phenotypes and the risk of diabetes mellitus in UK Biobank participants taking antidepressants and antipsychotics.

## Methods

### Sample and phenotype data

The UK Biobank data collection methods have been described previously in Bycroft et al (2018) and detailed study protocols are available online (http://www.ukbiobank.ac.uk/resources/ and http://biobank.ctsu.ox.ac.uk/crystal/docs.cgi/) (36,37). The study was approved by the North-West Research Ethics Committee (ref 06/MREC08/65). All participants provided written informed consent, and those who withdrew consent after providing their sample for genetic analysis were excluded from the data extraction. Data for 502,527 UK Biobank participants were considered in this study.

Participants for this study were selected based on the criteria of taking one or more psychotropic drugs. Participants were asked during a verbal interview if they were taking any ‘regular prescription medication’, and to provide the name of the medication if so. Both generic and proprietary names were recorded by UK Biobank. In these instances, we reviewed the alternative names for equivalent drugs and combined them under the generic name for analysis. For additional detail, please refer to the supplementary methods section and supplementary figure 1. We identified a sample of 44,051 participants taking a drug of interest for this study.

The UK Biobank measured a variety of biochemical markers in blood samples collected at the baseline visit. Glycated haemoglobin (HbA1c) was measured with the High Performance Liquid Chromatography (HPLC) method on a Bio-Rad VARIANT II Turbo analyser. The HbA1c analytical range was 15-184 mmol/mol and this measurement was recorded for over 92% of the UK Biobank cohort. Data on diabetes diagnosis (self-reported and confirmed by ICD-10 diagnosis when available), antidiabetic medications, CYP2D6 and/or CYP2C19 enzyme inhibitors and body mass index (BMI) were also downloaded. Further detail is available in the supplementary methods. We identified 49 individuals who reported taking antidiabetic medication but stated they do not have diabetes. They were excluded from the analysis due to uncertainty about their diagnosis. A total of 40,783 participants taking a psychotropic drug of interest also had HbA1c measurements available.

### Genetic data and quality control

The UK Biobank conducted genome-wide genotyping for 488,377 participants. Genotyping was performed using the Affymetrix UK BiLEVE Axiom array on an initial sample of 50,000 and the Affymetrix UK Biobank Axiom® array was used on all later participants (36). These arrays include over 820,000 variants (SNPs and indel markers) and have good coverage of pharmacogenetics variants. Quality control and imputation of over 90 million variants was performed by a collaborative group led by the Wellcome Trust Centre for Human Genetics (36). Fully imputed genetic data was downloaded in March 2018. Further local post-imputation quality control was performed in each ethnic group separately to remove variants with minor allele frequency below 1% and/or Fisher information score (a measure of the imputation accuracy for each SNP) of less than 0.3. Individuals with greater than 10% missingness, excessive genetic relatedness (greater than 10 third-degree relatives based on kinship calculations as provided centrally by UK Biobank) or mismatch between reported and genetically inferred sex were removed.

We included both European and non-European subjects in this analysis. A list of approximately 408,000 participants of European ancestry was provided centrally by UK Biobank, based on a combination of principal component analysis (PCA) and self-reported ethnicity data (36). Further local analysis was conducted to determine the genetic ancestry of the remaining participants: Two rounds of PCA were performed using the PC-AiR algorithm, and relatedness was estimated using PC-Relate (38–41). This resulted in the following groups: East Asian 0.5% (N=2,464), South Asian 2% (N=8,964), African 2% (N=9,233) or admixed with predominantly European origin 2.5% (N=11,251). A further 6,686 did not cluster with any main group and were excluded from analysis. One of each pair of participants with a kinship score greater than 0.083 (approximately third-degree relatives) were excluded from the analysis. This results in a total of 40,129 participants to exclude, across all ethnicities. After these quality control procedures, a total of 33,149 participants taking antidepressant and/or antipsychotic medication with HbA1c and good quality genetic data were included in the analysis. Please see supplementary figure 1 for a CONSORT diagram detailing these steps.

### Assigning CYP metabolic phenotype

We extracted regions of interest for each CYP2D6 and CYP2C19, defined as being one megabase (Mb) upstream of the 5’ end of the gene and one megabase downstream of the 3’ end of the gene (see supplementary table 1). Several of the SNPs of interest in this study (i.e., those that define either CYP2D6 or CYP2C19 star alleles) are rare (MAF < 0.01) and therefore fail standard quality control protocols. For rare SNPs of interest included on the genotype panel we used Evoker v2.4 to create intensity plots and performed visual checks to determine if the data for these SNPs was reliable enough to include (42). We reviewed a total of six genotyped SNPs for CYP2C19 and five for CYP2D6. SNPs with distinct allelic clusters were included in this study. For the rare, imputed SNPs, we included only those that met a higher Fisher information score threshold of 0.6. We reviewed a total of seven imputed SNPs for CYP2C19 and five for CYP2D6. These steps enabled the inclusion of an additional four relevant SNPs for CYP2C19, and three for CYP2D6. The extraction of data and identification of rare SNPs was conducted separately for each ancestry group.

Haplotypes for our sample were constructed based on extracted imputed genetic data using Beagle version 5.0 (43,44). An input map and reference panel from the 1,000 genome project were used (45). The phased data was used to construct haplotypes for all participants according to the star allele nomenclature system (20,46). We grouped individuals into CYP2C19 metabolic phenotype groups based on the activity of the individual haplotypes and resulting diplotypes (46). We grouped individuals into CYP2D6 metabolic phenotype groups according to the Gaedigk activity score method (47,48). Haplotypes containing no star-allele defining SNP variants were classified as wild-type (*1) alleles for the corresponding gene. Because not all star allele-defining SNPs were available in our genetic dataset, we expect a fraction of haplotypes to be misclassified as wild-type. Nonetheless, as the cumulative reported frequency of the missing SNPs is very low, we expect the number of misclassified haplotypes to be small.. In addition, we did not have data on CYP2D6 copy number variants (CNVs). This means we are not able to define CYP2D6 ultra-rapid metabolisers, or other whole gene deletions (e.g., CYP2D6*5).

### Statistical analysis

We conducted a grouped analysis of all tricyclic antidepressants, as previous evidence suggests that they all cause an increase in HbA1c to some extent (49). We did not analyse SSRIs as a group due to variable evidence on their influence on HbA1c in the literature (15,17,49). Any antidepressants taken by over 1,800 participants were analysed independently (amitriptyline, citalopram, fluoxetine, sertraline, paroxetine, venlafaxine). Medications were grouped according to whether their primary metabolic pathway was catalysed by CYP2D6 or CYP2C19, based on the Maudsley Prescribing Guidelines and CPIC guidelines (10,31,32). Tricyclic antidepressants that are known CYP2C19 substrates are: amitriptyline, clomipramine, doxepin, imipramine and trimipramine. SSRIs that are known CYP2C19 substrates are: citalopram, escitalopram, and sertraline. Tricyclic antidepressants that are known substrates for CYP2D6 include: amitriptyline, clomipramine, duloxetine, and doxepin. SSRIs that are known substrates for CYP2D6 are: fluoxetine, fluvoxamine, paroxetine, sertraline, as well as the SNRIs mirtazapine and venlafaxine (10,50).

No single antipsychotic drug had sufficient sample size to allow for individual analysis. Therefore, we included all antipsychotic drugs known to be metabolised at least in part by CYP2D6: aripiprazole, clozapine, fluphenazine, haloperidol, olanzapine, perphenazine, pimozide, risperidone, zuclopenthixol, thioridazine. CYP2C19 does not play a significant role in the metabolism of antipsychotics (10).

For each drug or drug group, we ran linear regression models with HbA1c as the outcome of interest and CYP450 metabolic phenotype and diabetes status as the main explanatory variables. All statistical models were adjusted to account for any participant taking antidiabetic treatment or taking drugs, psychotropic or otherwise, that are known inhibitors of the enzymes of interest. Additional covariates included were BMI, sex, age, and genetically determined ancestry group. We investigated the interaction of diabetes status and CYP metabolic phenotype. Where this interaction was significant (*p* < 0.05) we conducted a stratified analysis separating participants into two groups based on their diabetes status.

Some of these analyses are nested (individual drug analyses overlap with drug group analyses), and, as such, we concluded that a Bonferroni correction for multiple testing would be excessively stringent (51). Therefore, we report uncorrected *p* values in all text and tables, but as recommended by Li *et al* (2012) (52), we have an adjusted significance threshold of *p* < 0.05/2 = 0.025 (threshold for a suggestive association *p* < 0.1/2 = 0.05) for the two grouped analyses, and *p* < 0.05/6 = 0.0083 (threshold for a suggestive association *p* < 0.1/6 = 0.017) for the individual drug analyses examining six specific drugs. All statistical analyses were performed using R version 3.6.0 (53–55).

## Results

### Dataset

We identified 33,149 UK Biobank participants who reported taking at least one antidepressant or antipsychotic and had HbA1c and genetic data passing quality control (antidepressants N=31,579, antipsychotics N=2,699) (Table 1). Our sample included 22,632 (68.3%) females and 10,517 (31.7%) males (see Table 1). Mean age was 56.6±7.8 years, range 40 to 70 years. Full demographic data and summary statistics of our sample are shown in the Table 1 (see also supplementary Tables 5 and 6).

**Table 1.** Demographic data for study sample

|  | Antidepressants<br>(N=31579) | Antipsychotics<br>(N=2699) |
| --- | --- | --- |
| **CYP2D6 metabolic phenotype** |  |  |
| Normal metabolisers | 22486 (71.2%) | 1914 (70.9%) |
| Intermediate metabolisers | 7433 (23.5%) | 650 (24.1%) |
| Poor metabolisers | 1660 (5.3%) | 135 (5.0%) |
| **CYP2C19 metabolic phenotype** |  |  |
| Normal metabolisers | 12001 (38.0%) | 1004 (37.2%) |
| Intermediate metabolisers | 9367 (29.7%) | 789 (29.2%) |
| Poor metabolisers | 1065 (3.4%) | 100 (3.7%) |
| Rapid metabolisers | 7805 (24.7%) | 686 (25.4%) |
| Ultra-rapid metabolisers | 1341 (4.2%) | 120 (4.4%) |
| **Takes CYP2D6 inhibitors<sup>A</sup>** |  |  |
| No | 29713 (94.1%) | 2548 (94.4%) |
| Yes | 1866 (5.9%) | 151 (5.6%) |
| **Takes CYP2C19 inhibitors<sup>A</sup>** |  |  |
| No | 23608 (74.8%) | 2091 (77.5%) |
| Yes | 7971 (25.2%) | 608 (22.5%) |
| **Sex** |  |  |
| Female | 21752 (68.9%) | 1553 (57.5%) |
| Male | 9827 (31.1%) | 1146 (42.5%) |
| **Age** |  |  |
| Mean (SD) (years) | 56.6 (7.78) | 56.4 (8.12) |
| Range (median) (years) | 40-70 (58) | 40-70 (57) |
| **Ethnicity** |  |  |
| Caucasian | 29628 (93.8%) | 2403 (89.0%) |
| Admix Caucasian | 795 (2.5%) | 72 (2.7%) |
| African | 289 (0.9%) | 90 (3.3%) |
| East Asian | 43 (0.1%) | 12 (0.4%) |
| Other | 450 (1.4%) | 57 (2.1%) |
| South Asian | 374 (1.2%) | 65 (2.4%) |
| **Hb1Ac** |  |  |
| Mean (SD) (mmol/mol) | 37.1 (7.75) | 37.5 (8.31) |
| **Diabetes status** |  |  |
| No | 28776 (91.1%) | 2415 (89.5%) |
| Yes | 2803 (8.9%) | 284 (10.5%) |
| **Takes antidiabetic medications<sup>B</sup>** |  |  |
| No | 29573 (93.6%) | 2491 (92.3%) |
| Yes | 2006 (6.4%) | 208 (7.7%) |
| **BMI** |  |  |
| Mean (SD) (kg/m<sup>2</sup>) | 28.8 (5.66) | 29.1 (5.94) |

<sup>A</sup> CYP2C19 and CYP2D6 inhibitors identified through review of literature, including British National Formulary; <sup>B</sup> As defined by British National Formulary (59)

### Psychotropics prescribed in the UK Biobank

There were 28 different antidepressants identified in our sample (figure 1; supplementary Tables 7 and 8). Amitriptyline was the most common drug in our cohort (N=8,191). We identified 24 different antipsychotic drugs (figure 1; supplementary Table 9), with the most frequent antipsychotics being prochlorperazine (870 individuals, 30.9%), followed by olanzapine (499 individuals, 17.7%). Among UK Biobank participants taking antidepressants, 5.2% report taking more than one different antidepressant concurrently (of these, 2% report taking three or four). Of those taking antipsychotics, 4.5% report taking more than one different antipsychotic medication concurrently (of these, 7.4% report taking three or four). The co-prescription of an antidepressant with antipsychotics is very common, with 41.4% of subjects taking antipsychotics also taking at least one antidepressant.

**Figure 1.**
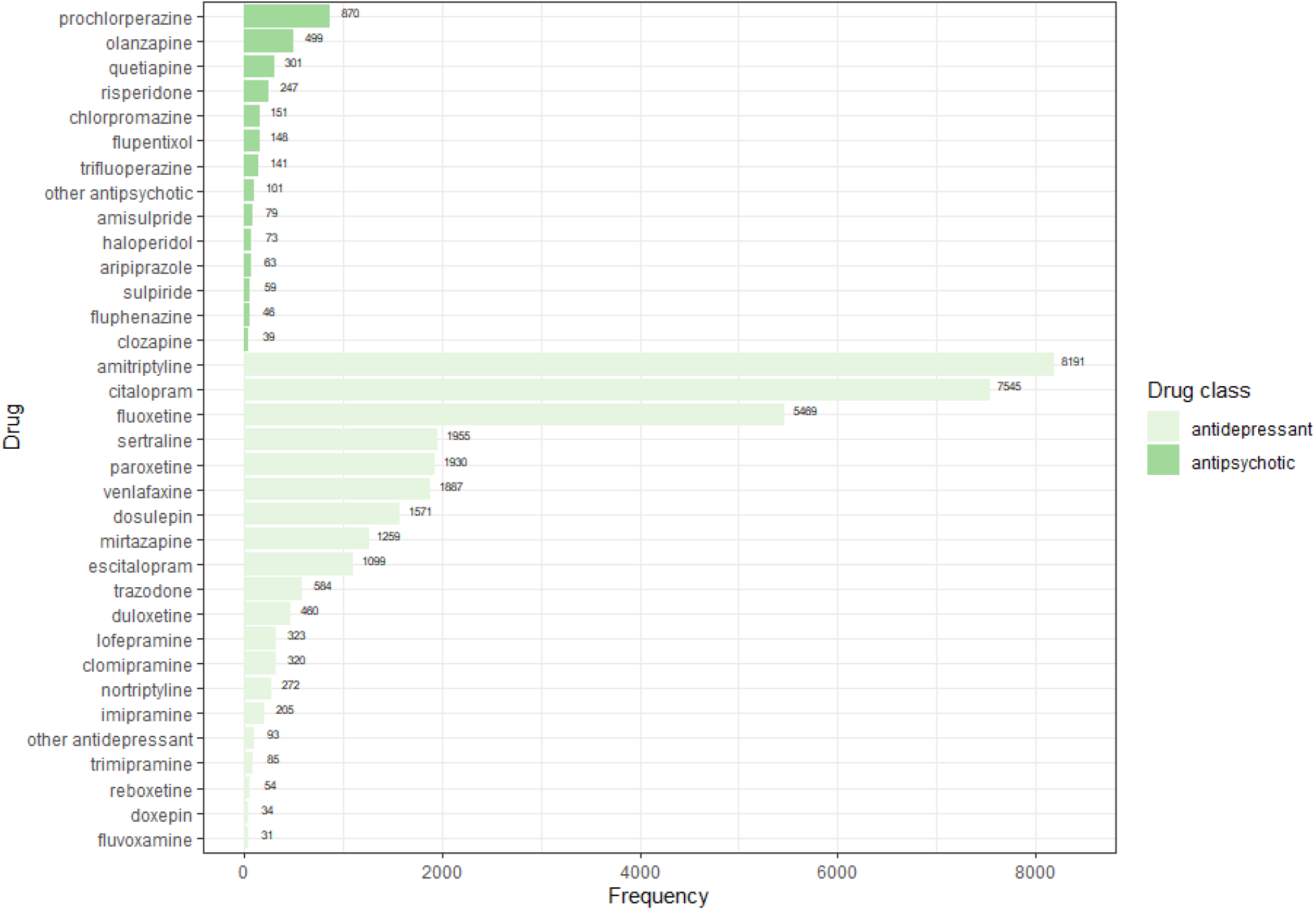
Frequency table of identified antipsychotics (blue bars) and antidepressants (green bars) in UK Biobank. **Other antipsychotic (N):** promazine (30), zuclopenthixol (25), perphenazine (12), pipotiazine (10), pericyazine (8), levopromazine (6), benperidol (4), pimozide (3), thioridazine (2), sertindole (1); **Other antidepressants (N)**: moclobemide (30), phenelzine (30), tranylcypromine (21), bupropion (6), mianserin (4), isocarboxazid (2).

The included covariates (diabetes status, antidiabetic medications, BMI, age, sex, and ethnicity) affected HbA1c as expected. Please refer to the supplementary methods for further details.

#### Antidepressants and CYP metabolic status

For several of the antidepressants investigated, we consistently found that the interaction of diabetes status and CYP2D6 and CYP2C19 metabolic phenotype is statistically significant (supplementary figure 2). Where this was the case, we stratified our analyses by whether participants had diabetes or not. Among all participants (regardless of diabetes status) taking paroxetine (SSRI), we observe significantly higher HbA1c levels among CPY2D6 poor metabolisers (mean difference: 2.43mmol/mol; 95% CI [1.23,3.63]; p = 7.77×10^−5^) (see table 2, figure 2, and supplementary table 10). A stratified analysis of diabetic participants taking fluoxetine (SSRI) reveals a suggestive association between CYP2D6 intermediate metabolisers and lower HbA1c levels compared to normal metabolisers (mean difference = -3.74mmol/mol; 95% CI [-6.82,-0.67]; p = 0.017) (see table 3, figure 2, and supplementary table 11). In participants taking venlafaxine (SNRI), we found that, amongst people with diabetes, poor metabolisers for CYP2D6 had higher HbA1c than normal metabolisers (mean difference: 10.15mmol/mol; 95% CI [2.63,17.67]; p = 0.008) (see table 4, figure 2, and supplementary table 12).

**Table 2.** Association between CYP2D6 metabolic phenotype and HbA1c levels among participants taking fluoxetine and paroxetine.

| Predictors | Paroxetine |  |  |  |
| --- | --- | --- | --- | --- |
|  | N | Estimates | CI | p |
| Diabetes | 174 | 6.85 | 5.11,8.59 | **<0.001** |
| CYP2D6 IM | 457 | 0.23 | -0.42,0.87 | 0.489 |
| CYP2D6 PM | 106 | 2.43 | 1.23,3.63 | **<0.001** |
| Observations | 1930 |  |  |  |
| R<sup>2</sup> / R<sup>2</sup> adjusted | 0.454 / 0.450 |  |  |  |

*Model adjusted by age, ethnicity, sex, taking inhibitors of CYP2D6, diabetes status, taking antidiabetics and BMI; Normal metabolisers of CYP2D6 taking paroxetine: 1,367*

**Table 3.** A) Association between CYP2D6 metabolic phenotype and HbA1c levels among participants taking fluoxetine; B) Stratified analysis of diabetes status among participants taking fluoxetine.

| Predictors | Fluoxetine |  |  |  |
| --- | --- | --- | --- | --- |
|  | N | Estimates | CI | p |
| Diabetes | 426 | 7.22 | 6.20,8.23 | **<0.001** |
| CYP2D6 IM | 1282 | 0.06 | -0.29,0.41 | 0.728 |
| CYP2D6 PM | 299 | 0.04 | -0.62,0.69 | 0.916 |
| Diabetes: CYP2D6 IM |  | -3.78 | -5.03,-2.53 | **<0.001** |
| Diabetes: CYP2D6 PM |  | -1.81 | -4.11,0.49 | 0.124 |
| Observations |  | 5469 |  |  |
| R<sup>2</sup> / R<sup>2</sup> adjusted |  | 0.467 / 0.465 |  |  |

**B**
| Diabetes |  |  |  |  | No diabetes |  |  |  |
| --- | --- | --- | --- | --- | --- | --- | --- | --- |
| Predictors | N | Estimates | CI | p | N | Estimates | CI | p |
| CYP2D6 IM | 100 | -3.74 | -6.82,-0.67 | 0.017 | 1182 | 0.05 | -0.21,0.31 | 0.696 |
| CYP2D6 PM | 24 | -0.94 | -6.61,4.73 | 0.745 | 275 | 0.04 | -0.43,0.52 | 0.859 |
| Observations |  |  | 426 |  |  | 5043 |  |  |
| R<sup>2</sup> / R<sup>2</sup> adjusted |  |  | 0.196 / 0.175 |  |  | 0.130 / 0.128 |  |  |

Model adjusted by age, ethnicity, sex, taking inhibitors of CYP2D6, taking antidiabetics and BMI; Normal metabolisers of CYP2D6: diabetes = 302

**Table 4.** A) Association between CYP2D6 metabolic phenotype and HbA1c levels among participants taking venlafaxine; B) Stratified analysis of diabetes status among participants taking venlafaxine.

| Venlafaxine |  |  |  |  |
| --- | --- | --- | --- | --- |
| Predictors | N | Estimates | CI | p |
| Diabetes | 182 | 5.68 | 4.04,7.33 | **1.77e-11** |
| CYP2D6 IM | 430 | -0.23 | -0.89,0.43 | 0.495 |
| CYP2D6 PM | 103 | -0.46 | -1.73,0.80 | 0.473 |
| Diabetes: CYP2D6 IM |  | 3.62 | 1.27,5.98 | **0.003** |
| Diabetes: CYP2D6 PM |  | 11.44 | 8.05,14.84 | **4.79e-11** |
| Observations |  |  | 1887 |  |
| R<sup>2</sup> / R<sup>2</sup> adjusted |  |  | 0.528 / 0.524 |  |

**B**
| Diabetes |  |  |  |  | No diabetes |  |  |  |
| --- | --- | --- | --- | --- | --- | --- | --- | --- |
| Predictors | N | Estimates | CI | p | N | Estimates | CI | p |
| CYP2D6 IM | 32 | 3.55 | -1.75,8.85 | 0.188 | 398 | -0.22 | -0.71,0.26 | 0.367 |
| CYP2D6 PM | 15 | 10.15 | 2.63,17.67 | **0.008** | 88 | -0.44 | -1.36,0.49 | 0.356 |
| Observations |  |  | 182 |  | 1703 |  |  |  |
| R<sup>2</sup> / R<sup>2</sup> adjusted |  |  | 0.280 / 0.233 |  | 0.122 / 0.116 |  |  |  |

Model adjusted by age, ethnicity, sex, taking inhibitors of CYP2D6, taking antidiabetics and BMI; Normal metabolisers of CYP2D6: diabetes = 135

**Figure 2.**
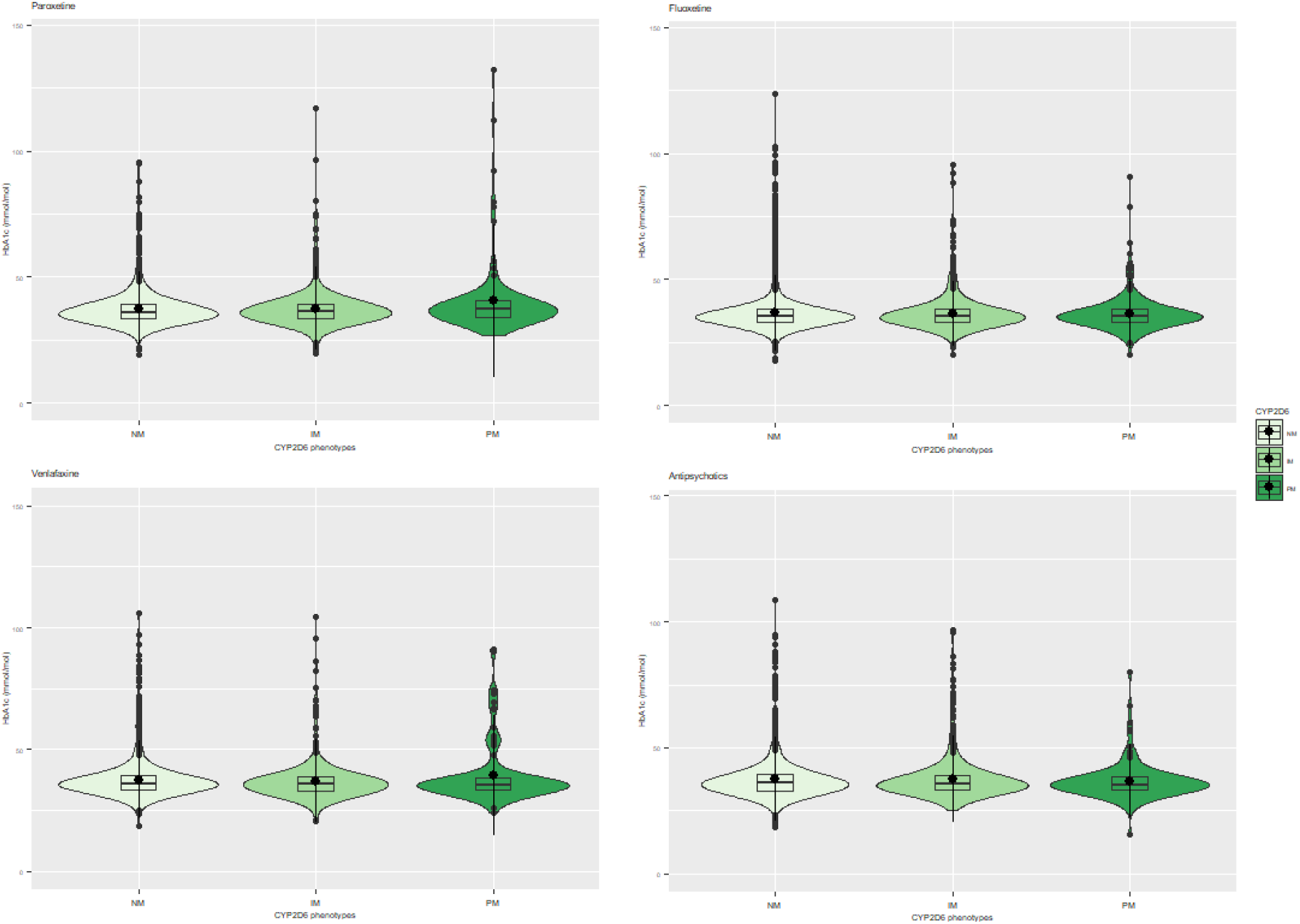
Violin plots showing the relationship between CYP2D6 metabolic status and HbA1c levels (mmol/mol) among subjects taking (from left to right) paroxetine, fluoxetine, venlafaxine, and all antipsychotics.

Stratified analyses of citalopram, sertraline, and amitriptyline did not reveal any significant association between the relevant CYP450 metabolic status and HbA1c levels (see supplementary tables 13-18).

Several tricyclic antidepressants were reported too infrequently to allow for single-drug analysis. Therefore, we grouped the remaining drugs of this class, excluding amitriptyline as its higher frequency would have heavily driven the findings. We again stratified the group based on diabetes status and found no significant associations between CYP2D6 or CYP2C19 derived metabolic groups and HbA1c (see supplementary Table 19 and 20).

We did not observe any significant association between HbA1c levels and CYP2C19 metabolic status in individuals taking antidepressants. In addition, we find that participants taking drugs that act as CYP2C19 inhibitors, regardless of CYP2C19 metabolic status, experience higher levels of HbA1c. Citalopram: mean difference: 0.36mmol/mol, 95% CI [0.07,0.65]; p = 0.016); Amitriptyline: mean difference: 0.37mmol/mol; 95% CI [0.09,0.64]; p = 0.009; Tricyclics: mean difference = 0.39mmol/mol; 95% CI [0.13,0.66]; p = 0.004). We did not see this relationship with sertraline (see supplementary Tables 13, 15, 17, 19).

### Antipsychotics and CYP metabolic status

We find no evidence that the metabolic phenotypes of CYP2D6 influence HbA1c levels amongst 2,699 people taking antipsychotic medications. Similarly, taking a CYP2D6 inhibitor drug was not significantly associated with HbA1c levels amongst people taking antipsychotic medication. See table 5, figure 2 and supplementary Table 21.

**Table 5.** Association between CYP2D6 metabolic phenotype and HbA1c levels in participants taking antipsychotics

| *Predictors* | *N/mean* | *HbA1c mmol/mol estimates* | *95% CI mmol/mol* | *p* |
| --- | --- | --- | --- | --- |
| CYP2D6 IM | 650 | -0.02 | -0.58,0.53 | 0.930 |
| CYP2D6 PM | 135 | -0.93 | -2.01,0.16 | 0.093 |
| Takes CYP2D6 inhibitor | 151 | 0.59 | -0.43,1.61 | 0.260 |
| Diabetes | 284 | 4.55 | 3.13,5.97 | **<0.001** |
| Observations | 2699 |  |  |  |
| R<sup>2</sup> / R<sup>2</sup> adjusted | 0.449 / 0.446 |  |  |  |

*Model adjusted by age, ethnicity, sex, taking inhibitors of CYP2D6, diabetes status, taking antidiabetics and BMI; Normal metabolisers of CYP2D6 = 1914.*

## Discussion

Non-normal metabolic phenotypes of CYP2D6 and CYP2C19 have been linked to QT prolongation (56,57), weight gain (58–61), hormonal changes among patients taking psychotropic medication, as well as increased risk of extrapyramidal adverse reactions to antipsychotics (62). However, recent studies and meta-analyses have yielded inconclusive or negative findings and the clinical significance of CYP450 metabolic phenotypes is still in question (30,63). Several studies agree that long-term antidepressant treatment increases risk of developing diabetes (4,64–66), but the extent to which this specific adverse drug reaction is impacted by genetics is unknown. To our knowledge, this study is the first to explore if variation in the CYP2D6 and CYP2C19 genes influences HbA1c levels in individuals taking antidepressants and antipsychotics. Most previous studies of CYP450 metabolic status and adverse drug reactions are limited by small sample sizes and low representation of the less common poor or ultra-rapid metabolisers (30,56–66). This study represents one of the largest available samples of individuals taking antidepressants and antipsychotics and includes a much higher number of extreme CYP450 metabolisers than seen in previous publications (N=9,878 non-wild-type CYP2D6 metabolisers and N=21,273 non-wild-type CYP2C19 metabolisers).

We find a significant association between CYP2D6 poor metabolisers and higher levels of HbA1c among all participants taking paroxetine with an average increase of 2.3mmol/mol, a substantial effect. The Clinical Pharmacogenetics Implementation Consortium (CPIC) guidelines recommend using lower doses of paroxetine for poor metabolisers of CYP2D6 (32). Thus, our findings are consistent with existing pharmacokinetic evidence and provide further support for the CPIC guidelines. Of interest, some research found that prolonged use of paroxetine was associated with phenocopying, an environmentally induced conversion of normal metabolisers to poor metabolisers (67–69).

We observe a significant interaction between diabetic status and non-wild-type CYP status for participants taking amitriptyline, fluoxetine, citalopram, sertraline, and venlafaxine. We conducted stratified analyses of these drugs and found suggestive evidence that, in diabetic participants taking venlafaxine, CYP2D6 poor and intermediate metabolisers have higher HbA1c levels. Like paroxetine, venlafaxine has been previously associated with an increased risk of diabetes (4,15,70). Our study finds that diabetic CYP2D6 poor metabolisers treated with venlafaxine have on average 10.15 mmol/mol higher HbA1c levels than diabetic normal metabolisers. Though this is a suggestive association only with a comparatively small sample size, it is consistent with the guidelines published by the Dutch Pharmacogenetics Working Group which suggest that CYP2D6 poor metabolisers should be treated with an alternative antidepressants or have their venlafaxine dose reduced (33). In addition, a stratified analysis reveals suggestive evidence that diabetic CYP2D6 intermediate metabolisers taking fluoxetine have lower HbA1c levels compared to diabetic CYP2D6 normal metabolisers. Although this is contrary to our initial hypothesis, there is some evidence to suggest that fluoxetine can lower HbA1c levels in diabetic patients, despite increasing risk of type 2 diabetes in non-diabetic patients (71–73). Our findings add support to this theory, suggesting that decreased CYP2D6 metabolism may in fact be somewhat beneficial for patients with diabetes who take fluoxetine.

Contrary to our hypotheses, we did not find evidence of associations between CYP2D6 or CYP2C19 metabolic status and HbA1c in people treated with amitriptyline and other tricyclics. Although CPIC guidelines exist for CYP2C19 and CYP2D6 poor metabolisers taking tricyclic antidepressants, they state that suggested dose alterations or treatment changes are optional based on the limited strength of existing evidence (31). Our analyses of tricyclics antidepressants and amitriptyline alone were adequately powered with over 400 poor metabolisers of each gene, making it one of the largest samples of abnormal CYP metabolisers available. However, the metabolic pathway of amitriptyline (and other tertiary amine tricyclic antidepressants) involves two steps: the first step is catalysed by CYP2C19 and produces an active metabolite (nortriptyline). The second step is the metabolism of nortriptyline to an inactive metabolite, via CYP2D6 (74,75). This is not adequately accounted for in this study, and thus future studies investigating the synergistic action of CYP2D6 and CYP2C19 on amitriptyline metabolism are required.

In addition, we did not find associations between CYP2D6 variation and HbA1c amongst people taking antipsychotics, nor did we observe an impact of CYP2D6 inhibitors. Given the total sample size of 2,699, we undertook a combined analysis including all antipsychotics, which have various levels of influence on glucose regulation and diabetes risk. Although this sample is the largest available with 135 CYP2D6 poor metabolisers overall, statistical power remains limited given the heterogeneity of the sample. Analysis in a larger sample would allow for the separate analysis of individual drugs and should yield more conclusive results. This limitation also applies to the less common antidepressants in our sample, which were included in grouped analyses only. Given that UK Biobank is a population study, utilizing existing data from large patient-based biobanks such as the Million Veteran Program could be a valuable continuation of this work (76). Biobanks from countries with more historically isolated populations, such as Finngen, may contain a higher proportion of some rare SNPs that are necessary to define additional CYP450 star alleles.

As well as being impacted by genetic variation, CYP2D6 and CYP2C19 enzyme activity is susceptible to inhibition by other compounds. We observed that taking CYP2C19 inhibitors (of which proton pump inhibitors were the most common in our sample) led to higher HbA1c levels in people taking tricyclic antidepressants, amitriptyline, and citalopram. Thus, based on our data, there is substantial potential for drug interactions and drug-drug-CYP2C19 interactions. These should be investigated further and considered for inclusion in future clinical guidelines. We did not find evidence that taking CYP2D6 inhibitors affected HbA1c levels in people taking antipsychotics or antidepressants. This enzyme inhibition could, however, still be important for other psychotropic adverse effects such as QT prolongation. It is also worth noting that CYP2C19 inhibitor drugs were taken by approximately a quarter of the subjects included in these analyses. That may have decreased the power to detect a significant association between the (genetically-determined) CYP2C19 metabolic status and HbA1c levels, because if the inhibitory impact of a drug was strong enough it would reduce or eliminate the impact of the genetic variation; i.e. an CYP2C19 normal metaboliser taking an inhibitor may have the same enzymatic activity and as CYP2C19 poor metaboliser taking an inhibitor. Future analyses with larger sample sizes should investigate this interaction further.

A clear limitation of this study is the reliance on certain self-reported data (including diabetes diagnosis). In addition, we have used only the baseline, cross-sectional UK Biobank data and therefore lack detail on treatment dose and duration. Most adverse drug reactions to antidepressants and antipsychotics are dose-dependent, and thus further analysis including this data is warranted. Besides, diabetes is a complex disease with many genetic and environmental risk factors. Although the SNP-based heritability of diabetes is estimated to be less than 20%, the inclusion of polygenic risk scores for diabetes may improve analyses of pharmacogenetic associations by capturing background genetic disease risk (77). A genome-wide gene-environment interaction study may also highlight other genes of potential interest. Finally, although we included participants of all ethnicities in this analysis, UK Biobank is predominantly European. There is a great deal of variation in the frequency of functional variants within the CYP450 genes across different populations (22,78), as well as in the risk of diabetes. The field of pharmacogenetics would be greatly benefitted by further study in more diverse samples.

Although both arrays used by UK Biobank have relatively good coverage of CYP2C19 and CYP2D6, several SNPs that define known star alleles were neither genotyped nor imputed or did not otherwise meet the criteria for inclusion as described in the methods. Therefore, we expect a small number of individuals to be misclassified as normal metabolisers. However, we anticipate this number to be small given the low minor allele frequency of the missing variants. We were unable to include CYP2D6 ultra-rapid metabolisers in this study, as copy number and other structural variants were not defined. CYP2D6 ultra-rapid metabolisers are the least common phenotypic group across all populations, with a frequency of less than 2% in European, South Asian, East Asian and Admixed European groups, and approximately 3-6% in African ancestry groups (22,78). CYP2D6 ultra-rapid metabolisers therefore represent a very small minority in our sample, and they have been combined with the normal metabolisers group by default. We estimate this to have a small effect on our results as we would expect ultra-rapid metabolisers to be less susceptible to adverse drug reactions, though it will be important to consider this group in future studies of treatment failure. The availability of whole genome sequencing data will improve the accuracy with which highly polymorphic pharmacogenes like CYP2D6 can be characterised, whilst still capturing the important splicing or non-coding variants that may be missed with exome sequencing data (79).

Overall, our findings are broadly consistent with existing guidelines for antidepressants and point towards the necessity to include more antidepressants and antipsychotics in pharmacogenetic clinical trials and experimental medicine studies. These results also suggest that there is a need for randomised double-blinded clinical trials to further explore genetic testing as a guide to antidepressant/antipsychotic treatment. Indeed, studies show that pharmacogenetic testing is practical (80), accurately predicts the outcomes of antidepressant treatments (81) and improves outcomes (82,83). It has also been demonstrated that it can reduce the total cost of antipsychotic treatment by 28% (84). Findings from this study need to be followed up with further longitudinal testing, with a focus on singular antidepressants and antipsychotics, more adverse drug reactions, and in more diverse populations.

## Data Availability

Data used in this paper is available to all UK Biobank approved researchers.

## Acknowledgements

The authors thank all UK Biobank participants.

This research has been conducted using the UK Biobank under application ID 20737 (PI: Andrew McQuillin, Co-I: Elvira Bramon). The UK Biobank study was approved by the North-West Research Ethics Committee (ref 06/MREC08/65) in accordance with the Declaration of Helsinki. This study was supported by a Medical Research Council doctoral studentships awarded to Isabelle Austin-Zimmerman, Anjali Bhat, and Jasmine Harju-Seppänen. Baihan Wang was supported by the China Scholarship Council-University College London Joint Research Scholarship. Haritz Irizar has received funding from the European Union’s Horizon 220 research and innovation programme under the Marie Sklodowska-Curie grant agreement no 747429 and is currently supported by a grant from the National Institute of Allergy and Infectious Diseases, National Institutes of Health. Gazaleh Fatemifar is funded by the American Heart Association Institutional Data Fellowship Program (AHA Award 17IF3389000). Elvira Bramon has recieved the following grant funding that supported this work: National Institute of Health Research UK (NIHR200756); Mental Health Research UK John Grace QC Scholarship 2018; BMA Margaret Temple Fellowship 2016; Medical Research Council New Investigator and Centenary Awards (G0901310, G1100583), MRC (G1100583); NIHR Biomedical Research Centre at University College London Hospitals NHS Foundation Trust and University College London.

## Notes

### Competing Interest Statement

The authors have declared no competing interest.

### Author Declarations

The UK Biobank study was approved by the North-West Research Ethics Committee (ref 06/MREC08/65) in accordance with the Declaration of Helsinki.

### Summary of Updates

Correcting author list and adding additional figure (fig2).

