## Supplementary material for "The influence of CYP2D6 and CYP2C19 genetic variation on diabetes mellitus risk in people taking antidepressants and antipsychotics"

### Methodology: Sample

The medication data recorded in the UK Biobank was collected through a verbal interview with a trained nurse. Participants were asked if they were currently taking any regular prescription medication. If they answered yes, the nurse asked them to detail medication they were taking. Dose, formulation or duration of treatment were not recorded, and any short-term medication (i.e., one week course of antibiotics) were not included.

A psychotropic drug was defined as any drug indicated, either officially licenced or through routine clinical practice, for the treatment of psychosis (schizophrenia), depression or bipolar disorder. Anxiolytic agents were not included in this study. The drugs of interest were identified through review of the British National Formulary (BNF) Drug Dictionary[1]. The list was expanded to include drugs licenced overseas and drugs no longer listed in the BNF. As the medication data in the UK Biobank are self-reported, it was necessary to identify all potential names (brand names and generic names in foreign languages) of the medications of interest. These alternate names were identified through review of published drug labels, the websites of the European Medicines Agency (EMA) [2], the Food and Drug Administration (FDA)[3], the Mayo Clinic[4], the independent website www.drugs.com[5] and the National Library of Medicine[6].

[1] MedicinesComplete - British National Formulary. https://www.medicinescomplete.com/mc/bnf/current/
[2] European Medicines Agency. https://www.ema.europa.eu/
[3] U S Food and Drug Administration. https://www.fda.gov/
[4] Mayo Clinic. https://www.mayoclinic.org/
[5] Drugs.com | Prescription Drug Information, Interactions & Side Effects. https://www.drugs.com/
[6] National Library of Medicine's LactMed Database. https://toxnet.nlm.nih.gov/newtoxnet/lactmed.htm
[7] Adapted from: CONSORT 2010 Statement, BMJ 2010;340:c332 http://www.consort-statement.org/

**Supplementary figure 1. Adapted CONSORT 2010 statement[7]**

CONSORT
TRANSPARENT REPORTING of TRIALS

**CONSORT 2010 Flow Diagram**

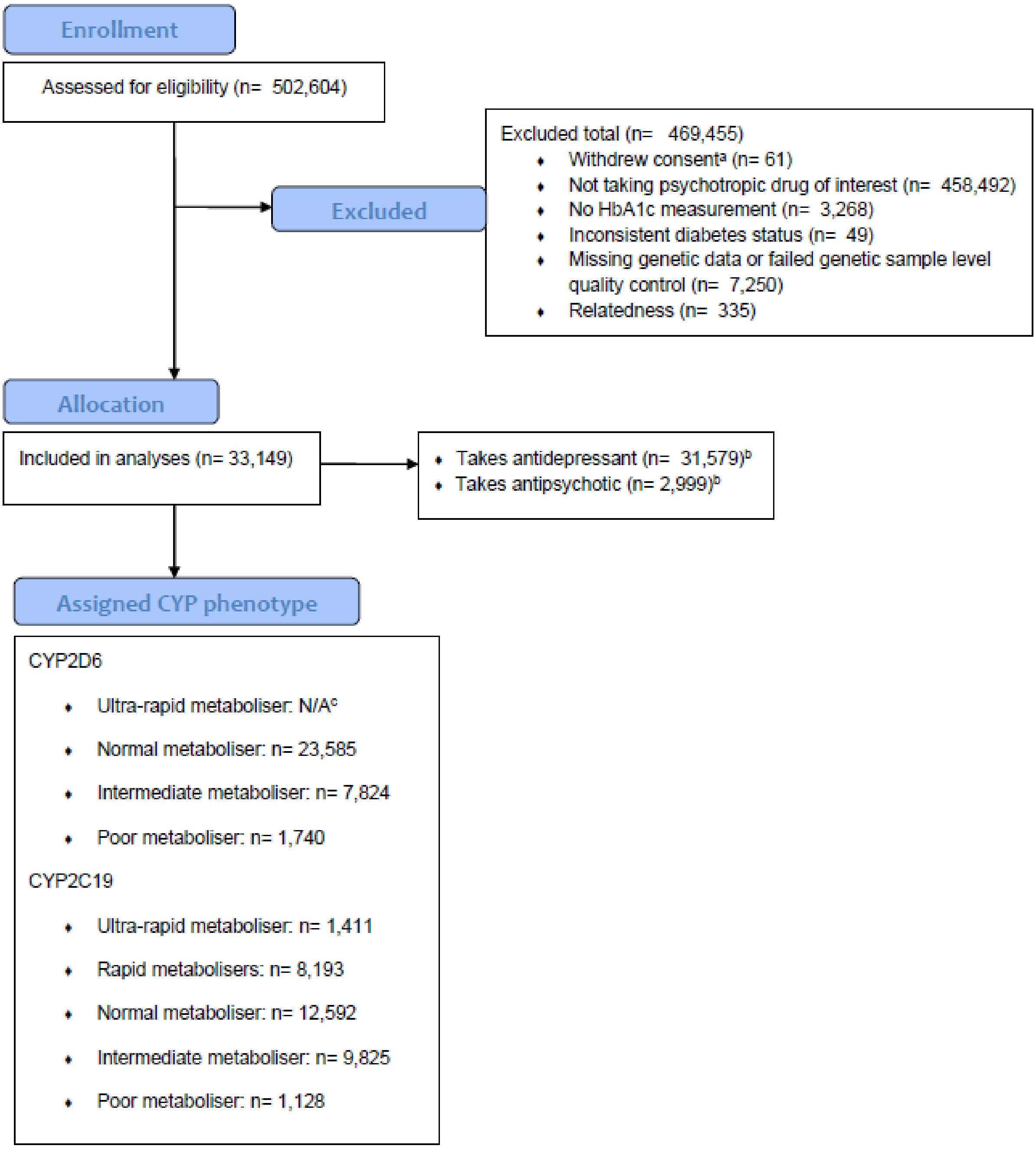


*[a] Number of participants who withdrew consent after the initial download of all phenotype data and before the analysis was conducted. Participants who withdrew consent prior to initial download of data were already removed; [b] Note that several subjects report taking both antidepressant(s) and antipsychotic(s) and are included in both analyses; [c] See main body of text for detail on why CYP2D6 ultra-rapid metabolisers were not defined in this analysis.*

**Methodology: Covariates**

### Diabetes

There were several items in the UK Biobank which could act as a source of information for whether the patient had diabetes or not, namely ICD-10 diagnosis, self-reported diagnosis of diabetes or self-reported use of antidiabetic medications. Though this would be the most reliable item to use for the purpose of our research, the ICD-10 data was incomplete, covering 410,316 participants and recording 3399 (0.82%) cases of diabetes mellitus. Based on self-reported data, the prevalence of diabetes mellitus in the UK Biobank was 5.28%. This is consistent with UK epidemiological studies which report the prevalence of diabetes mellitus at 7%[7]. We thus concluded that the self-reported data was more reliable in this instance. Based on the data available we were unable to differentiate between type 1 and type 2 diabetes.

### Anti-diabetic medications

We identified participants taking insulin, metformin, thiazolidinediones, sulfonylureas, meglitinides, alpha-glucosidase inhibitors. There were no participants within our dataset who have been taking glucagon-like peptide-1 or gastric inhibitory peptide as well as gliflozins. We also identified all brand names of anti-diabetic medications and converted these to their generic equivalents and created a dichotomous variable which reflected the use of the antidiabetic medications.

### BMI

A high BMI is an independent risk factor for diabetes; hence it was included in our analyses. BMI at baseline was downloaded directly from the UK Biobank.

### Enzyme inhibitors

We identified any participants taking drugs that inhibit CYP2D6 and CYP2C19 activity[8]. In the sample, 1,969 participants were taking a CYP2D6 inhibitor drug including: ranitidine, celecoxib, metoclopramide, chlorphenamine, terbinafine, hydroxyzine or promethazine. We also identified 8,340 participants taking drugs that are known CYP2C19 inhibitors including omeprazole, esomeprazole, lansoprazole, pantoprazole, oestrogen, cimetidine, modafinil, topiramate, indomethacin or oxcarbazepine.

[7] Whicher, C. A., O'Neill, S., & Holt, R. I. G. (2020). Diabetes in the UK: 2019. *Diabetic Medicine*, *37* (2), 242–247. https://doi.org/10.1111/dme.14225

[8] *CYTOCHROME P450 DRUG INTERACTION TABLE - Drug Interactions*. (n.d.). Retrieved September 1, 2020, from https://drug-interactions.medicine.iu.edu/MainTable.asp

**Covariates in antidepressants' model**

As expected, in every antidepressants' model, having diabetes, taking antidiabetic medications, raising BMI and increasing age was associated with higher HbA1c (all p <0.001). In all models apart from venlafaxine, South Asian ethnicity was associated with higher HbA1c level (p range <0.001 - 0.050). African ethnicity was associated with higher HbA1c in following models: citalopram, paroxetine, venlafaxine (p range <0.001 – 0.005). Admix Caucasian ethnicity was associated with higher HbA1c levels in fluoxetine models (p = 0.006 and p = 0.027 accordingly) and Other ethnicity in tricyclic and amitriptyline models (p <0.001, p = 0.002). Male sex was associated with higher HbA1c in citalopram and fluoxetine models. Please refer to supplementary tables 10, 11, 14, 16, 17, 18, 21.

Methods: Assigning metabolic phenotype

We extracted regions of interest for each CYP2D6 and CYP2C19, defined as being 1 megabase (Mb) either side of each gene. Start and stop coordinates are provided in supplementary table 1 and were identified using the University of California Santa Cruz Human Genome Browser[9].

**Supplementary table 1. Showing start and stop positions used to extract relevant genetic data from full UKB sample.**

| GENE (CHROMOSOME, POSITION) | START POSITION FOR DATA EXTRACTION | STOP POSITION FOR DATA EXTRACTION |
|---|---|---|
| **CYP2C19 (CHR10: 96447882- 96612671)** | 95447882 | 97612671 |
| **CYP2D6 (CHR 22: 42522501- 42526883)** | 41522501 | 43526883 |

**Supplementary table 2. Frequencies of CYP2C19 diplotypes and metabolic phenotypes in 33,149 Biobank participants taking antidepressants or antipsychotics.**

| Diplotype | Frequency (N) | Percentage (%) |
|---|---|---|
| **Poor metaboliser** | | |
| CYP2C19*1 / CYP2C19*1 | 10270 | 30.98 |
| CYP2C19*1 / CYP2C19*13 | 1080 | 3.26 |
| CYP2C19*13 / CYP2C19*1 | 1034 | 3.12 |
| CYP2C19*13 / CYP2C19*13 | 104 | 0.31 |
| **Intermediate metaboliser** | | |

[9] Kent W, Sugnet C, Furey T, et al. The Human Genome Browser at UCSC. *Genome Res*. 2002;12 (6):996-1006.

| | | |
|---|---|---|
| CYP2C19*1 / CYP2C19*2 | 2843 | 8.58 |
| CYP2C19*2 / CYP2C19*1 | 2766 | 8.34 |
| CYP2C19*17 / CYP2C19*2 | 1048 | 3.16 |
| CYP2C19*2 / CYP2C19*17 | 969 | 2.92 |
| CYP2C19*1 / CYP2C19*3 | 475 | 1.43 |
| CYP2C19*3 / CYP2C19*1 | 448 | 1.35 |
| CYP2C19*13 / CYP2C19*2 | 333 | 1 |
| CYP2C19*2 / CYP2C19*13 | 289 | 0.87 |
| CYP2C19*17 / CYP2C19*3 | 183 | 0.55 |
| CYP2C19*3 / CYP2C19*17 | 166 | 0.5 |
| **Normal metaboliser** | | |
| CYP2C19*2 / CYP2C19*2 | 773 | 2.33 |
| CYP2C19*3 / CYP2C19*2 | 152 | 0.46 |
| CYP2C19*2 / CYP2C19*3 | 125 | 0.38 |
| **Rapid metaboliser** | | |
| CYP2C19*1 / CYP2C19*17 | 3755 | 11.33 |
| CYP2C19*17 / CYP2C19*1 | 3667 | 11.06 |
| CYP2C19*13 / CYP2C19*17 | 382 | 1.15 |
| CYP2C19*17 / CYP2C19*13 | 357 | 01.08 |
| **Ultra-rapid metaboliser** | | |
| CYP2C19*17 / CYP2C19*17 | 1411 | 4.26 |

‘/’ distinguishes two homologous chromosomes. ‘;’ connects several different alleles on the same chromosome

**Supplementary table 3. Frequencies of CYP2D6 diplotypes and metabolic phenotypes in 33,149 Biobank participants taking antidepressants or antipsychotics.**

| Diplotype | Frequency (N) | Percentage (%) |
|---|---|---|
| **Normal metaboliser** | | |

| | | |
|---|---|---|
| CYP2D6*29 / CYP2D6*29 | 5716 | 17.24 |
| CYP2D6*1 / CYP2D6*29 | 2848 | 8.59 |
| CYP2D6*29 / CYP2D6*1 | 2751 | 8.3 |
| CYP2D6*1 / CYP2D6*1 | 2049 | 6.18 |
| CYP2D6*4 / CYP2D6*1 | 1556 | 4.69 |
| CYP2D6*1 / CYP2D6*4 | 1533 | 4.62 |
| CYP2D6*29 / CYP2D6*41 | 1157 | 3.49 |
| CYP2D6*41 / CYP2D6*29 | 1155 | 3.48 |
| CYP2D6*41 / CYP2D6*1 | 853 | 2.57 |
| CYP2D6*1 / CYP2D6*41 | 815 | 2.46 |
| CYP2D6*29 / CYP2D6*9;CYP2D6*29 | 416 | 1.25 |
| CYP2D6*9;CYP2D6*29 / CYP2D6*29 | 408 | 1.23 |
| CYP2D6*41 / CYP2D6*41 | 337 | 1.02 |
| CYP2D6*1 / CYP2D6*9;CYP2D6*29 | 208 | 0.63 |
| CYP2D6*9;CYP2D6*29 / CYP2D6*1 | 186 | 0.56 |
| CYP2D6*10 / CYP2D6*29 | 158 | 0.48 |
| CYP2D6*3A / CYP2D6*1 | 132 | 0.4 |
| CYP2D6*29 / CYP2D6*10 | 129 | 0.39 |
| CYP2D6*1 / CYP2D6*3A | 118 | 0.36 |
| **Intermediate metaboliser** | | |
| CYP2D6*4 / CYP2D6*29 | 2512 | 7.58 |
| CYP2D6*29 / CYP2D6*4 | 2331 | 7.03 |
| CYP2D6*41 / CYP2D6*4 | 611 | 1.84 |
| CYP2D6*4 / CYP2D6*41 | 589 | 1.78 |
| CYP2D6*29 / CYP2D6*3A | 246 | 0.74 |
| CYP2D6*3A / CYP2D6*29 | 244 | 0.74 |

| | | |
|---|---|---|
| CYP2D6*10 / CYP2D6*4 | 222 | 0.67 |
| CYP2D6*4 / CYP2D6*10 | 219 | 0.66 |
| CYP2D6*4 / CYP2D6*9;CYP2D6*29 | 169 | 0.51 |
| CYP2D6*9;CYP2D6*29 / CYP2D6*4 | 140 | 0.42 |
| **Poor metaboliser** | | |
| CYP2D6*4 / CYP2D6*4 | 1468 | 4.43 |
| CYP2D6*3A / CYP2D6*4 | 103 | 0.31 |
| CYP2D6*4 / CYP2D6*3A | 95 | 0.29 |

'/' distinguishes two homologous chromosomes. ';' connects several different alleles on the same chromosome

**Supplementary table 4. HbA1c levels and CYP phenotypes across individual and groups of medications.**

| | **CYP2D6** | | | **CYP2C19** | | | | |
|---|---|---|---|---|---|---|---|---|
| | PM | IM | NM | PM | IM | NM | RM | UM |
| **Models** | HbA1c *[mmol/mol]**, (SD) | | | | | | | |
| Antipsychotics | 36.78 (7.26) | 37.60 (8.54) | 37.53 (8.30) | - | - | - | - | - |
| Tricyclics | 37.60 (7.79) | 37.79 (8.38) | 37.86 (8.21) | 37.75 (7.80) | 37.6 (7.73) | 37.88 (8.30) | 38.00 (8.81) | 38.13 (7.88) |
| Amitriptyline | 37.52 (7.86) | 37.93 (8.60) | 37.85 (8.18) | 37.61 (7.28) | 37.64 (7.84) | 37.92 (8.31) | 38.00 (8.86) | 38.08 (8.06) |
| Fluoxetine | 36.50 (6.90) | 36.47 (6.50) | 36.65 (7.50) | - | - | - | - | - |
| Paroxetine | 40.46 (15.05) | 37.50 (8.27) | 37.38 (7.27) | - | - | - | - | - |
| Citalopram | - | - | - | 36.89 (7.54) | 36.48 (6.90) | 36.67 (7.56) | 36.51 (7.23) | 36.01 (6.16) |
| Sertraline | - | - | - | 35.30 (4.35) | 36.99 (7.24) | 37.15 (7.43) | 37.13 (7.37) | 37.04 (8.50) |

| | | | | | | | | |
|---|---|---|---|---|---|---|---|---|
| Venlafaxine | 39.49 (12.38) | 37.13 (8.56) | 37.58 (8.06) | - | - | - | - | - |

* HbA1c levels diagnostic for impaired glucose regulation: normal < 42 mmol/mol, prediabetes 42 - 47 mmol/mol, diabetes ≥48 mmol/mol

**Supplementary table 5. Characteristics of CYP2C19 metabolic phenotype in our sample**

| | NM (N=12592) | IM (N=9825) | PM (N=1128) | RM (N=8193) | UM (N=1411) | Overall (N=33149) |
|---|---|---|---|---|---|---|
| **Age (years)** | | | | | | |
| Mean (SD) | 56.6 (7.79) | 56.6 (7.85) | 56.2 (8.01) | 56.7 (7.72) | 56.6 (7.80) | 56.6 (7.80) |
| **Sex** | | | | | | |
| Female | 8623 (68.5%) | 6751 (68.7%) | 749 (66.4%) | 5547 (67.7%) | 962 (68.2%) | 22632 (68.3%) |
| Male | 3969 (31.5%) | 3074 (31.3%) | 379 (33.6%) | 2646 (32.3%) | 449 (31.8%) | 10517 (31.7%) |
| **Ethnicity** | | | | | | |
| European | 11762 (93.4%) | 9205 (93.7%) | 1062 (94.1%) | 7670 (93.6%) | 1307 (92.6%) | 31006 (93.5%) |
| Admix European | 342 (2.7%) | 223 (2.3%) | 27 (2.4%) | 198 (2.4%) | 46 (3.3%) | 836 (2.5%) |
| African | 127 (1.0%) | 110 (1.1%) | 10 (0.9%) | 95 (1.2%) | 16 (1.1%) | 358 (1.1%) |
| East Asian | 25 (0.2%) | 13 (0.1%) | 1 (0.1%) | 12 (0.1%) | 0 (0%) | 51 (0.2%) |
| Other | 178 (1.4%) | 149 (1.5%) | 19 (1.7%) | 109 (1.3%) | 24 (1.7%) | 479 (1.4%) |
| South Asian | 158 (1.3%) | 125 (1.3%) | 9 (0.8%) | 109 (1.3%) | 18 (1.3%) | 419 (1.3%) |
| **HbA1c (mmol/mol)** | | | | | | |
| Mean (SD) | 37.2 (7.89) | 37.0 (7.44) | 37.0 (7.02) | 37.2 (7.90) | 37.1 (7.84) | 37.1 (7.73) |
| **Diabetes** | | | | | | |
| Yes | 1117 (8.9%) | 827 (8.4%) | 105 (9.3%) | 762 (9.3%) | 125 (8.9%) | 2936 (8.9%) |
| No | 11475 (91.1%) | 8998 (91.6%) | 1023 (90.7%) | 7431 (90.7%) | 1286 (91.1%) | 30213 (91.1%) |
| **Taking antidiabetic medication** | | | | | | |
| Yes | 819 (6.5%) | 580 (5.9%) | 67 (5.9%) | 532 (6.5%) | 100 (7.1%) | 2098 (6.3%) |

| | | | | | | |
|---|---|---|---|---|---|---|
| No | 11773 (93.5%) | 9245 (94.1%) | 1061 (94.1%) | 7661 (93.5%) | 1311 (92.9%) | 31051 (93.7%) |
| **BMI** | | | | | | |
| Mean (SD) | 28.8 (5.67) | 28.7 (5.69) | 28.6 (5.34) | 28.7 (5.66) | 28.6 (5.73) | 28.7 (5.67) |
| **Taking CYP2C19 inhibitor** | | | | | | |
| Yes | 3184 (25.3%) | 2364 (24.1%) | 283 (25.1%) | 2068 (25.2%) | 360 (25.5%) | 8259 (24.9%) |
| No | 9408 (74.7%) | 7461 (75.9%) | 845 (74.9%) | 6125 (74.8%) | 1051 (74.5%) | 24890 (75.1%) |

**Supplementary table 6. Characteristics of CYP2D6 metabolic phenotype in our sample**

| | NM (N=23585) | IM (N=7824) | PM (N=1740) | Overall (N=33149) |
|---|---|---|---|---|
| **Age (years)** | | | | |
| Mean (SD) | 56.6 (7.80) | 56.6 (7.82) | 56.6 (7.71) | 56.6 (7.80) |
| Median [Min, Max] | 58.0 [40.0, 71.0] | 58.0 [40.0, 70.0] | 58.0 [40.0, 70.0] | 58.0 [40.0, 71.0] |
| **Sex** | | | | |
| Female | 16086 (68.2%) | 5355 (68.4%) | 1191 (68.4%) | 22632 (68.3%) |
| Male | 7499 (31.8%) | 2469 (31.6%) | 549 (31.6%) | 10517 (31.7%) |
| **Ethnicity** | | | | |
| Caucasian | 22027 (93.4%) | 7342 (93.8%) | 1637 (94.1%) | 31006 (93.5%) |
| Admix Caucasian | 620 (2.6%) | 181 (2.3%) | 35 (2.0%) | 836 (2.5%) |
| African | 262 (1.1%) | 78 (1.0%) | 18 (1.0%) | 358 (1.1%) |
| East Asian | 36 (0.2%) | 10 (0.1%) | 5 (0.3%) | 51 (0.2%) |
| Other | 346 (1.5%) | 108 (1.4%) | 25 (1.4%) | 479 (1.4%) |
| South Asian | 294 (1.2%) | 105 (1.3%) | 20 (1.1%) | 419 (1.3%) |
| **HbA1c (mmol/mol)** | | | | |
| Mean (SD) | 37.1 (7.74) | 37.0 (7.57) | 37.3 (8.34) | 37.1 (7.73) |
| **Diabetes** | | | | |
| Yes | 2106 (8.9%) | 670 (8.6%) | 160 (9.2%) | 2936 (8.9%) |
| No | 21479 (91.1%) | 7154 (91.4%) | 1580 (90.8%) | 30213 (91.1%) |
| **Taking antidiabetic medication** | | | | |

| | | | | |
|---|---|---|---|---|
| Yes | 1488 (6.3%) | 487 (6.2%) | 123 (7.1%) | 2098 (6.3%) |
| No | 22097 (93.7%) | 7337 (93.8%) | 1617 (92.9%) | 31051 (93.7%) |
| **BMI (kg/m²)** | | | | |
| Mean (SD) | 28.7 (5.65) | 28.8 (5.67) | 28.8 (5.80) | 28.7 (5.67) |
| **Taking CYP2D6 inhibitor** | | | | |
| Yes | 1401 (5.9%) | 440 (5.6%) | 102 (5.9%) | 1943 (5.9%) |
| No | 22184 (94.1%) | 7384 (94.4%) | 1638 (94.1%) | 31206 (94.1%) |

**Supplementary table 7. CYP2D6 metabolic phenotypes of people taking antidepressants**

| | NM (N=23749) | IM (N=7816) | PM (N=1757) | Overall (N=33367) |
|---|---|---|---|---|
| Antidepressant | 23794 (100%) | 7816 (100%) | 1757 (100%) | 33367 (100%) |
| **Tricyclic** | | | | |
| amitriptyline | 5840 (24.5%) | 1929 (24.7%) | 422 (24.0%) | 8191 (24.5%) |
| dosulepin | 1158 (4.9%) | 349 (4.5%) | 64 (3.6%) | 1571 (4.7%) |
| lofepramine | 231 (1.0%) | 71 (0.9%) | 21 (1.2%) | 323 (1.0%) |
| clomipramine | 221 (0.9%) | 91 (1.2%) | 8 (0.5%) | 320 (1.0%) |
| nortriptyline | 197 (0.8%) | 63 (0.8%) | 12 (0.7%) | 272 (0.8%) |
| imipramine | 139 (0.6%) | 51 (0.7%) | 15 (0.9%) | 205 (0.6%) |
| trimipramine | 62 (0.3%) | 19 (0.2%) | 4 (0.2%) | 85 (0.3%) |
| **SSRI** | | | | |
| citalopram | 5381 (22.6%) | 1753 (22.4%) | 411 (23.4%) | 7545 (22.6%) |
| fluoxetine | 3888 (16.3%) | 1282 (16.4%) | 299 (17.0%) | 5469 (16.4%) |
| sertraline | 1394 (5.9%) | 456 (5.8%) | 105 (6.0%) | 1955 (5.9%) |
| paroxetine | 1367 (5.7%) | 457 (5.8%) | 106 (6.0%) | 1930 (5.8%) |
| escitalopram | 795 (3.3%) | 249 (3.2%) | 55 (3.1%) | 1099 (3.3%) |
| **SNRI** | | | | |
| venlafaxine | 1354 (5.7%) | 430 (5.5%) | 103 (5.9%) | 1887 (5.7%) |
| duloxetine | 325 (1.4%) | 119 (1.5%) | 16 (0.9%) | 460 (1.4%) |
| **Tetracyclic** | | | | |
| mirtazapine | 869 (3.7%) | 316 (4.0%) | 74 (4.2%) | 1259 (3.8%) |

| | | | | |
|---|---|---|---|---|
| **SARI** | | | | |
| trazodone | 412 (1.7%) | 139 (1.8%) | 33 (1.9%) | 584 (1.8%) |
| **NRI** | | | | |
| reboxetine | 38 (0.2%) | 10 (0.1%) | 6 (0.3%) | 54 (0.2%) |
| **OTHER** | | | | |
| other | 123 (0.5%) | 32 (0.4%) | 3 (0.2%) | 158 (0.5%) |

NM - normal metaboliser, PM - poor metaboliser, IM – intermediate metaboliser, SSRI – Selective Serotonin Reuptake Inhibitor, SNRI – Selective Noradrenaline Reuptake Inhibitor, MOI – Monoamine Oxidase Inhibitor, NRI – Noradrenaline Reuptake Inhibitor, NDRI – Noradrenaline Dopamine Reuptake Inhibitor, SARI – Serotonin Antagonist and Reuptake Inhibitor

Other - doxepin, fluvoxamine, phenelzine, moclobemide, tranylcypromine, bupropion, mianserin, isocarboxazid

**Supplementary table 8. CYP2C19 metabolic phenotypes of people taking antidepressants**

| | NM (N=12689) | IM (N=9889) | PM (N=1122) | RM (N=8241) | UM (N=1426) | Overall (N=33367) |
|---|---|---|---|---|---|---|
| Antidepressant | 12689 (100%) | 9889 (100%) | 1122 (100%) | 8241 (100%) | 1426 (100%) | 33367 (100%) |
| **Tricyclic** | | | | | | |
| amitriptyline | 3116 (24.6%) | 2483 (25.1%) | 263 (23.4%) | 1961 (23.8%) | 368 (25.8%) | 8191 (24.5%) |
| dosulepin | 603 (4.8%) | 439 (4.4%) | 60 (5.3%) | 388 (4.7%) | 81 (5.7%) | 1571 (4.7%) |
| lofepramine | 127 (1.0%) | 95 (1.0%) | 4 (0.4%) | 82 (1.0%) | 15 (1.1%) | 323 (1.0%) |
| clomipramine | 126 (1.0%) | 96 (1.0%) | 12 (1.1%) | 75 (0.9%) | 11 (0.8%) | 320 (1.0%) |
| nortriptyline | 98 (0.8%) | 73 (0.7%) | 7 (0.6%) | 85 (1.0%) | 9 (0.6%) | 272 (0.8%) |
| imipramine | 74 (0.6%) | 59 (0.6%) | 5 (0.4%) | 59 (0.7%) | 8 (0.6%) | 205 (0.6%) |
| trimipramine | 38 (0.3%) | 26 (0.3%) | 2 (0.2%) | 13 (0.2%) | 6 (0.4%) | 85 (0.3%) |
| **SSRI** | | | | | | |
| citalopram | 2923 (23.0%) | 2205 (22.3%) | 232 (20.7%) | 1882 (22.8%) | 303 (21.2%) | 7545 (22.6%) |
| fluoxetine | 2065 (16.3%) | 1642 (16.6%) | 195 (17.4%) | 1338 (16.2%) | 229 (16.1%) | 5469 (16.4%) |
| sertraline | 760 (6.0%) | 587 (5.9%) | 67 (6.0%) | 465 (5.6%) | 76 (5.3%) | 1955 (5.9%) |
| paroxetine | 731 (5.8%) | 580 (5.9%) | 79 (7.0%) | 478 (5.8%) | 62 (4.3%) | 1930 (5.8%) |

| | | | | | | |
|---|---|---|---|---|---|---|
| escitalopram | 396 (3.1%) | 345 (3.5%) | 39 (3.5%) | 263 (3.2%) | 56 (3.9%) | 1099 (3.3%) |
| **SNRI** | | | | | | |
| venlafaxine | 717 (5.7%) | 536 (5.4%) | 71 (6.3%) | 483 (5.9%) | 80 (5.6%) | 1887 (5.7%) |
| duloxetine | 167 (1.3%) | 143 (1.4%) | 20 (1.8%) | 105 (1.3%) | 25 (1.8%) | 460 (1.4%) |
| **Tetracyclic** | | | | | | |
| mirtazapine | 439 (3.5%) | 359 (3.6%) | 40 (3.6%) | 356 (4.3%) | 65 (4.6%) | 1259 (3.8%) |
| **SARI** | | | | | | |
| trazodone | 220 (1.7%) | 169 (1.7%) | 18 (1.6%) | 155 (1.9%) | 22 (1.5%) | 584 (1.8%) |
| **NRI** | | | | | | |
| reboxetine | 21 (0.2%) | 15 (0.2%) | 2 (0.2%) | 13 (0.2%) | 3 (0.2%) | 54 (0.2%) |
| **OTHER** | | | | | | |
| other | 68 (0.5%) | 37 (0.4%) | 6 (0.5%) | 40 (0.5%) | 7 (0.5%) | 158 (0.5%) |

NM - normal metaboliser, PM - poor metaboliser, IM – intermediate metaboliser, RM – rapid metaboliser, UM – ultra-rapid metaboliser, SSRI – Selective Serotonin Reuptake Inhibitor, SNRI – Selective Noradrenaline Reuptake Inhibitor, MOI – Monoamine Oxidase Inhibitor, NRI – Noradrenaline Reuptake Inhibitor, NDRI – Noradrenaline Dopamine Reuptake Inhibitor, SARI – Serotonin Antagonist and Reuptake Inhibitor, Other - doxepin, fluvoxamine, phenelzine, moclobemide, tranylcypromine, bupropion, mianserin, isocarboxazid

**Supplementary table 9. CYP2D6 metabolic phenotype of individuals taking antipsychotics**

| | NM (N=2004) | IM (N=671) | PM (N=142) | Overall (N=2817) |
|---|---|---|---|---|
| Antipsychotic | 2004 (100%) | 671 (100%) | 142 (100%) | 2817 (100%) |
| **Medication** | | | | |
| prochlorperazine | 607 (30.3%) | 221 (32.9%) | 42 (29.6%) | 870 (30.9%) |
| olanzapine | 352 (17.6%) | 114 (17.0%) | 33 (23.2%) | 499 (17.7%) |
| quetiapine | 215 (10.7%) | 74 (11.0%) | 12 (8.5%) | 301 (10.7%) |
| risperidone | 181 (9.0%) | 56 (8.3%) | 10 (7.0%) | 247 (8.8%) |
| chlorpromazine | 105 (5.2%) | 40 (6.0%) | 6 (4.2%) | 151 (5.4%) |
| flupentixol | 107 (5.3%) | 36 (5.4%) | 5 (3.5%) | 148 (5.3%) |
| trifluoperazine | 110 (5.5%) | 25 (3.7%) | 6 (4.2%) | 141 (5.0%) |
| amisulpride | 57 (2.8%) | 17 (2.5%) | 5 (3.5%) | 79 (2.8%) |

| | | | | |
|---|---|---|---|---|
| haloperidol | 51 (2.5%) | 17 (2.5%) | 5 (3.5%) | 73 (2.6%) |
| aripiprazole | 43 (2.1%) | 16 (2.4%) | 4 (2.8%) | 63 (2.2%) |
| sulpiride | 41 (2.0%) | 14 (2.1%) | 4 (2.8%) | 59 (2.1%) |
| other | 135 (6.7%) | 41 (6.1%) | 10 (7.0%) | 186 (6.6%) |

NM - normal metaboliser, PM - poor metaboliser, IM – intermediate metaboliser. Other - fluphenazine, clozapine, promazine, zuclopenthxol, perphenazine, pipotiazine, periciazine, levomepromazine, benperidol, pimozide, thioridazine, sertindole

**Supplementary figure 2. Interaction between diabetes status and metabolic phenotypes among subjects taking, from left to right, (a) tricyclic antidepressants; (b) Amitriptyline; (c) Fluoxetine; (d) Venlafaxine; (e) Citalopram; (f) Sertraline.**

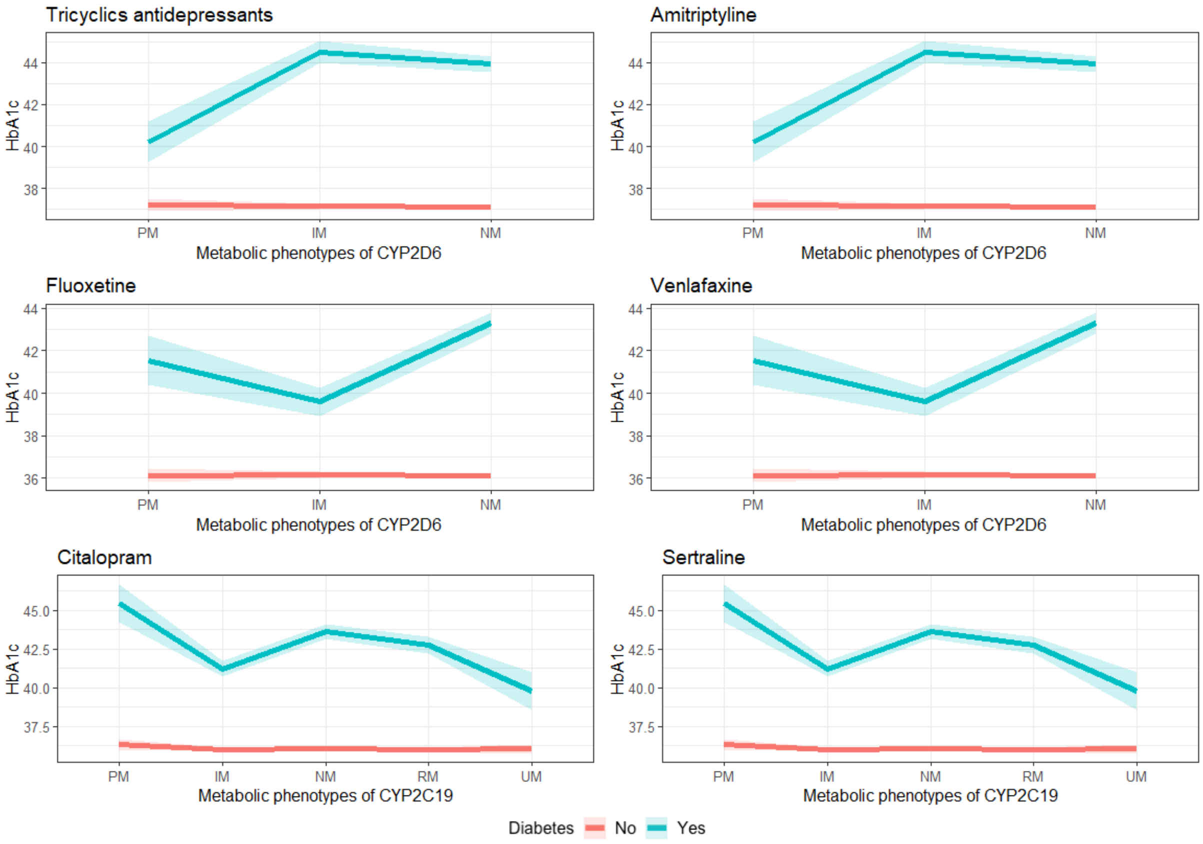


**Supplementary table 10. Association between CYP2D6 metabolic phenotype and HbA1c within individuals taking paroxetine – additional detail**

| | Paroxetine | | |
|---|---|---|---|
| *Predictors* | *Estimates* | *CI* | *p* |
| CYP2D6 IM | 0.23 | -0.42,0.87 | 0.489 |
| CYP2D6 PM | 2.43 | 1.23,3.63 | **<0.001** |

| | | | |
|---|---|---|---|
| Takes CYP2D6 inhibitor | -0.34 | -1.55,0.87 | 0.577 |
| Sex: Male | 0.41 | -0.16,0.98 | 0.158 |
| Age at recruitment | 0.15 | 0.11,0.19 | **<0.001** |
| Ethnicity: Admix Caucasian | 0.14 | -1.39,1.68 | 0.856 |
| Ethnicity: African | 6.41 | 2.96,9.86 | **<0.001** |
| Ethnicity: East Asian | -0.90 | -12.83,11.03 | 0.882 |
| Ethnicity: Other | 0.56 | -1.94,3.06 | 0.661 |
| Ethnicity: South Asian | 6.76 | 3.92,9.59 | **<0.001** |
| Diabetes | 6.85 | 5.11,8.59 | **<0.001** |
| BMI | 0.14 | 0.09,0.19 | **<0.001** |
| Antidiabetics | 12.89 | 10.88,14.89 | **<0.001** |
| Observations | 1930 | | |
| $R^2$ / $R^2$ adjusted | 0.454 / 0.450 | | |

**Supplementary table 11. Association between CYP2D6 metabolic phenotype and HbA1c within individuals taking fluoxetine – additional detail**

| | **Fluoxetine** | | |
|---|---|---|---|
| *Predictors* | *Estimates* | *CI* | *p* |
| CYP2D6 IM | 0.06 | -0.29,0.41 | 0.728 |
| CYP2D6 PM | 0.04 | -0.62,0.69 | 0.916 |
| Diabetes: CYP2D6 IM | -3.78 | -5.03,-2.53 | **<0.001** |
| Diabetes: CYP2D6 PM | -1.81 | -4.11,0.49 | 0.124 |
| Takes CYP2D6 inhibitor | 0.01 | -0.62,0.64 | 0.970 |
| Sex: Male | 0.36 | 0.04,0.67 | **0.027** |
| Age at recruitment | 0.15 | 0.13,0.17 | **<0.001** |
| Ethnicity: Admix Caucasian | 1.27 | 0.36,2.17 | **0.006** |

| | | | |
|---|---|---|---|
| Ethnicity: African | 0.80 | -0.75,2.34 | 0.314 |
| Ethnicity: East Asian | 1.84 | -1.63,5.31 | 0.299 |
| Ethnicity: Other | 0.28 | -0.91,1.47 | 0.640 |
| Ethnicity: South Asian | 3.68 | 2.03,5.34 | **<0.001** |
| Diabetes | 7.22 | 6.20,8.23 | **<0.001** |
| BMI | 0.16 | 0.13,0.18 | **<0.001** |
| Antidiabetics | 12.50 | 11.39,13.62 | **<0.001** |
| Observations | 5469 | | |
| $R^2$ / $R^2$ adjusted | 0.467 / 0.465 | | |

**Supplementary table 12. Association between CYP2D6 metabolic phenotype and HbA1c within individuals taking venlafaxine – additional detail**

| | **Venlafaxine** | | |
|---|---|---|---|
| *Predictors* | *Estimates* | *CI* | *p* |
| CYP2D6 IM | -0.23 | -0.89,0.43 | 0.489 |
| CYP2D6 PM | -0.46 | -1.73,0.80 | 0.495 |
| Diabetes: CYP2D6 IM | 3.62 | 1.27,5.98 | **0.003** |
| Diabetes: CYP2D6 PM | 11.44 | 8.05,14.84 | **4.79e-11** |
| Takes CYP2D6 inhibitor | -0.37 | -1.50,0.77 | 0.525 |
| Sex: Male | 0.41 | -0.14,0.97 | 0.146 |
| Age at recruitment | 0.14 | 0.11,0.18 | **<0.001** |
| Ethnicity: Admix Caucasian | 0.42 | -1.17,2.02 | 0.602 |
| Ethnicity: African | 4.59 | 1.37,7.80 | **0.005** |
| Ethnicity: East Asian | -0.96 | -12.42,10.51 | 0.870 |
| Ethnicity: Other | 1.97 | -0.62,4.56 | 0.136 |
| Ethnicity: South Asian | 2.63 | -0.09,5.35 | 0.058 |

| | | | |
|---|---|---|---|
| Diabetes | 5.68 | 4.04,7.33 | **1.77e-11** |
| BMI | 0.15 | 0.10,0.20 | **<0.001** |
| Antidiabetics | 14.82 | 12.97,16.67 | **<0.001** |
| Observations | 1885 | | |
| $R^2$ / $R^2$ adjusted | 0.528 / 0.524 | | |

Model adjusted by age, ethnicity, sex, taking inhibitors of CYP2D6, taking antidiabetics and BMI

Normal metabolisers of CYP2D6: 1,352; normal metabolisers of CYP2D6:diabetes = 135

**Supplementary table 13. Association between CYP2C19 metabolic phenotype and HbA1c within individuals taking citalopram**

| | Citalopram | | |
|---|---|---|---|
| *Predictors* | *Estimates* | *CI* | *p* |
| CYP2C19 IM | -0.06 | -0.36,0.24 | 0.701 |
| CYP2C19 PM | 0.25 | -0.48,0.99 | 0.500 |
| CYP2C19 RM | -0.07 | -0.39,0.24 | 0.650 |
| CYP2C19 UM | -0.04 | -0.68,0.61 | 0.913 |
| CYP2C19 IM : Diabetes | -2.33 | -3.41,-1.25 | **<0.001** |
| CYP2C19 PM : Diabetes | 1.62 | -0.97,4.21 | 0.221 |
| CYP2C19 RM : Diabetes | -0.76 | -1.92,0.40 | 0.198 |
| CYP2C19 UM : Diabetes | -3.78 | -6.28,-1.28 | **0.003** |
| Takes CYP2C19 inhibitor | 0.36 | 0.07,0.65 | **0.016** |
| Sex: Male | 0.29 | 0.02,0.55 | **0.032** |
| Age at recruitment | 0.13 | 0.12,0.15 | **<0.001** |
| Ethnicity: Admix Caucasian | 0.02 | -0.75,0.80 | 0.958 |
| Ethnicity: African | 1.90 | 0.58,3.22 | **0.005** |
| Ethnicity: East Asian | 0.45 | -2.83,3.73 | 0.788 |
| Ethnicity: Other | 0.81 | -0.16,1.78 | 0.100 |

| | | | |
|---|---|---|---|
| Ethnicity: South Asian | 3.80 | 2.78,4.81 | **<0.001** |
| Diabetes | 7.53 | 6.55,8.51 | **<0.001** |
| BMI | 0.16 | 0.14,0.19 | **<0.001** |
| Antidiabetics | 12.59 | 11.65,13.52 | **<0.001** |
| Observations | | 7545 | |
| $R^2$ / $R^2$ adjusted | | 0.470 / 0.468 | |

**Supplementary table 14. Stratified analysis of people taking citalopram**

| | Citalopram | | | | | | | |
|---|---|---|---|---|---|---|---|---|
| | Diabetes | | | | No diabetes | | | |
| *Predictors* | *N* | *Estimates* | *CI* | *p* | *N* | *Estimates* | *CI* | *p* |
| CYP2C19 IM | 181 | -2.42 | -4.99, 0.16 | 0.066 | 2024 | -2.42 | -0.29, 0.18 | 0.635 |
| CYP2C19 PM | 19 | 1.37 | -4.81, 7.54 | 0.664 | 213 | 1.37 | -0.31, 0.80 | 0.392 |
| CYP2C19 RM | 140 | -1.03 | -3.82, 1.76 | 0.470 | 1742 | -1.03 | -0.31, 0.17 | 0.557 |
| CYP2C19 UM | 20 | -4.07 | -10.09, 1.94 | 0.184 | 283 | -4.07 | -0.52, 0.46 | 0.894 |
| Observations | | | 583 | | | 6962 | | |
| $R^2$ / $R^2$adjusted | | | 0.189 / 0.170 | | | 0.127 / 0.125 | | |

*Model adjusted by age, ethnicity, sex, taking inhibitors of CYP2D6, taking antidiabetics and BMI*

*Normal metabolisers of CYP2C19: citalopram diabetes = 223, sertraline diabetes = 74*

**Supplementary table 15. Association between CYP2C19 metabolic phenotype and HbA1c within individuals taking sertraline**

| | Sertraline | | |
|---|---|---|---|
| *Predictors* | *Estimates* | *CI* | *p* |
| CYP2C19 IM | 0.13 | -0.49,0.76 | 0.679 |

| | | | |
|---|---|---|---|
| CYP2C19 PM | -0.58 | -2.02,0.86 | 0.429 |
| CYP2C19 RM | -0.17 | -0.84,0.50 | 0.618 |
| CYP2C19 UM | -0.47 | -1.82,0.89 | 0.500 |
| CYP2C19 IM : Diabetes | -0.64 | -2.68,1.40 | 0.539 |
| CYP2C19 PM : Diabetes | -5.84 | -11.12,-0.56 | **0.030** |
| CYP2C19 RM : Diabetes | 0.17 | -1.96,2.30 | 0.876 |
| CYP2C19 UM : Diabetes | 8.52 | 3.31,13.73 | **0.001** |
| Takes CYP2C19 inhibitor | -0.05 | -0.62,0.52 | 0.860 |
| Sex: Male | 0.04 | -0.49,0.58 | 0.876 |
| Age at recruitment | 0.13 | 0.10,0.16 | **<0.001** |
| Ethnicity: Admix Caucasian | -0.37 | -1.71,0.97 | 0.588 |
| Ethnicity: African | 2.31 | -0.51,5.13 | 0.108 |
| Ethnicity: East Asian | 2.91 | -2.51,8.32 | 0.293 |
| Ethnicity: Other | -0.46 | -2.61,1.68 | 0.671 |
| Ethnicity: South Asian | 2.29 | 0.00,4.57 | **0.050** |
| Diabetes | 5.80 | 4.01,7.60 | **<0.001** |
| BMI | 0.18 | 0.13,0.22 | **<0.001** |
| Antidiabetics | 11.57 | 9.84,13.30 | **<0.001** |
| Observations | | 1955 | |
| $R^2$ / $R^2$ adjusted | | 0.438 / 0.433 | |

**Supplementary table 16. Stratified analysis of people taking sertraline**

| | | | | | | | | |
|---|---|---|---|---|---|---|---|---|
| | | | **Sertraline** | | | | | |
| | **Diabetes** | | | | **No diabetes** | | | |
| *Predictors* | *N* | *Estimates* | *CI* | *p* | *N* | *Estimates* | *CI* | *p* |

| | | | | | | | | |
|---|---|---|---|---|---|---|---|---|
| CYP2C19 IM | 54 | -2.42 | -5.07, 3.85 | 0.787 | 533 | -2.42 | -0.35, 0.59 | 0.621 |
| CYP2C19 PM | 5 | 1.37 | -20.28, 3.49 | 0.165 | 62 | 1.37 | -1.66, 0.52 | 0.306 |
| CYP2C19 RM | 47 | -1.03 | -4.28, 5.10 | 0.863 | 418 | -1.03 | -0.69, 0.33 | 0.494 |
| CYP2C19 UM | 71 | -4.07 | -3.85, 19.04 | 0.192 | 71 | -4.07 | -1.53, 0.52 | 0.330 |
| Observations | | | 185 | | | 1770 | | |
| $R^2$ / $R^2$adjusted | | | 0.232 / 0.174 | | | 0.133 / 0.127 | | |

Model adjusted by age, ethnicity, sex, taking inhibitors of CYP2D6, taking antidiabetics and BMI

Normal metabolisers of CYP2C19: citalopram diabetes = 223, sertraline diabetes = 74

**Supplementary table 17. Association between CYP2D6 and CYP2C19 metabolic phenotype and HbA1c within Amitriptyline**

| | **Amitriptyline** | | |
|---|---|---|---|
| *Predictors* | *Estimates* | *CI* | *p* |
| CYP2D6 IM | 0.04 | -0.28,0.37 | 0.789 |
| CYP2D6 PM | 0.10 | -0.51,0.72 | 0.740 |
| CYP2C19 IM | -0.10 | -0.43,0.23 | 0.545 |
| CYP2C19 PM | 0.01 | -0.79,0.81 | 0.978 |
| CYP2C19 RM | -0.13 | -0.49,0.23 | 0.476 |
| CYP2C19 UM | 0.02 | -0.66,0.70 | 0.951 |
| Diabetes : CYP2D6 IM | 1.03 | 0.02,2.03 | **0.046** |
| Diabetes: CYP2D6 PM | -3.41 | -5.44,-1.38 | **0.001** |
| CYP2C19 IM : Diabetes | -0.07 | -1.12,0.97 | 0.889 |
| CYP2C19 PM : Diabetes | -1.67 | -4.01,0.67 | 0.163 |
| CYP2C19 RM : Diabetes | 0.46 | -0.61,1.53 | 0.401 |

| | | | |
|---|---|---|---|
| CYP2C19 UM : Diabetes | 0.06 | -2.06,2.18 | 0.955 |
| Takes CYP2D6 inhibitor | -0.28 | -0.78,0.23 | 0.280 |
| Takes CYP2C19 inhibitor | 0.37 | 0.09,0.65 | **0.009** |
| Sex: Male | 0.09 | -0.20,0.38 | 0.532 |
| Age at recruitment | 0.12 | 0.10,0.14 | **<0.001** |
| Ethnicity: Admix Caucasian | 0.07 | -0.84,0.98 | 0.886 |
| Ethnicity: African | 2.57 | 1.42,3.72 | **<0.001** |
| Ethnicity: East Asian | -1.41 | -5.83,3.00 | 0.531 |
| Ethnicity: Other | 1.74 | 0.66,2.82 | **0.002** |
| Ethnicity: South Asian | 2.90 | 1.81,4.00 | **<0.001** |
| Diabetes | 6.68 | 5.64,7.73 | **<0.001** |
| Antidiabetics | 12.36 | 11.43,13.29 | **<0.001** |
| BMI | 0.16 | 0.13,0.18 | **<0.001** |
| Observations | 8191 | | |

**Supplementary table 18. Stratified analysis of people taking amitriptyline**

| | | | | | | | | |
|---|---|---|---|---|---|---|---|---|
| | **Amitriptyline** | | | | | | | |
| | **Diabetes** | | | | **No diabetes** | | | |
| *Predictors* | *N* | *Estimates* | *CI* | *p* | *N* | *Estimates* | *CI* | *p* |
| CYP2D6 IM | 197 | 1.18 | -0.93, 3.30 | 0.273 | 1732 | 0.04 | -0.19, 0.28 | 0.733 |
| CYP2D6 PM | 39 | -2.92 | -7.23, 1.39 | 0.184 | 383 | 0.10 | -0.35, 0.55 | 0.671 |
| CYP2C19 IM | 248 | -0.26 | -2.46, 1.95 | 0.819 | 2235 | -0.09 | -0.33, 0.15 | 0.473 |
| CYP2C19 PM | 31 | -1.43 | -6.33, 3.48 | 0.568 | 232 | 0.04 | -0.54, 0.62 | 0.902 |

| | | | | | | | | |
|---|---|---|---|---|---|---|---|---|
| CYP2C19 RM | 235 | 0.07 | -2.19, 2.32 | 0.952 | 1726 | -0.12 | -0.38, 0.14 | 0.366 |
| CYP2C19 UM | 38 | -0.09 | -4.55, 4.37 | 0.969 | 330 | 0.05 | -0.45, 0.54 | 0.851 |
| Observations | | | 874 | | | 7317 | | |
| $R^2$ / $R^2$ adjusted | | | 0.158 / 0.142 | | | 0.100 / 0.098 | | |

**Supplementary table 19. Association between CYP2D6 and CYP2C19 metabolic phenotype and HbA1c within individuals taking tricyclic antidepressants**

| | Tricyclic antidepressants | | |
|---|---|---|---|
| *Predictors* | *Estimates* | *CI* | *p* |
| CYP2D6 IM | 0.04 | -0.26,0.34 | 0.793 |
| CYP2D6 PM | 0.13 | -0.46,0.72 | 0.660 |
| CYP2C19 IM | -0.11 | -0.42,0.20 | 0.495 |
| CYP2C19 PM | -0.07 | -0.83,0.69 | 0.857 |
| CYP2C19 RM | -0.09 | -0.43,0.24 | 0.594 |
| CYP2C19 UM | 0.12 | -0.52,0.77 | 0.709 |
| Diabetes : CYP2D6 IM | 0.53 | -0.43,1.50 | 0.279 |
| Diabetes: CYP2D6 PM | -3.85 | -5.76,-1.95 | **<0.001** |
| CYP2C19 IM : Diabetes | -0.49 | -1.48,0.51 | 0.338 |
| CYP2C19 PM : Diabetes | -1.34 | -3.50,0.81 | 0.222 |
| CYP2C19 RM : Diabetes | 0.27 | -0.75,1.29 | 0.604 |
| CYP2C19 UM : Diabetes | -0.52 | -2.52,1.49 | 0.613 |
| Takes CYP2D6 inhibitor | -0.26 | -0.73,0.21 | 0.279 |
| Takes CYP2C19 inhibitor | 0.39 | 0.13,0.66 | **0.004** |
| Sex: Male | 0.11 | -0.16,0.38 | 0.428 |

| | | | |
|---|---|---|---|
| Age at recruitment | 0.12 | 0.11,0.14 | **<0.001** |
| Ethnicity: Admix Caucasian | 0.18 | -0.67,1.03 | 0.686 |
| Ethnicity: African | 2.47 | 1.36,3.57 | **<0.001** |
| Ethnicity: East Asian | 0.81 | -2.55,4.16 | 0.638 |
| Ethnicity: Other | 2.19 | 1.16,3.22 | **<0.001** |
| Ethnicity: South Asian | 2.55 | 1.52,3.59 | **<0.001** |
| Diabetes | 6.98 | 5.99,7.97 | **<0.001** |
| BMI | 0.16 | 0.13,0.18 | **<0.001** |
| Antidiabetics | 12.45 | 11.58,13.33 | **<0.001** |
| Observations | 9095 | | |
| $R^2$ / $R^2$ adjusted | 0.484 / 0.482 | | |

**Supplementary table 20. Stratified analysis of people taking tricyclic antidepressants**

| | | | | | | | | |
|---|---|---|---|---|---|---|---|---|
| | | | | **Tricyclics** | | | | |
| | | **Diabetes** | | | | **No diabetes** | | |
| *Predictors* | *N* | *Estimates* | *CI* | *p* | *N* | *Estimates* | *CI* | *p* |
| CYP2D6 IM | 1949 | 0.73 | -1.33, 2.78 | 0.488 | 208 | 0.04 | -0.18, 0.26 | 0.740 |
| CYP2D6 PM | 419 | -3.30 | -7.36, 0.76 | 0.111 | 44 | 0.12 | -0.31, 0.54 | 0.596 |
| CYP2C19 IM | 2475 | -0.61 | -2.73, 1.50 | 0.570 | 274 | -0.09 | -0.32, 0.13 | 0.414 |
| CYP2C19 PM | 252 | -1.26 | -5.77, 3.25 | 0.584 | 37 | -0.04 | -0.59, 0.52 | 0.899 |
| CYP2C19 RM | 1940 | -0.08 | -2.24, 2.09 | 0.945 | 257 | -0.08 | -0.33, 0.16 | 0.501 |
| CYP2C19 UM | 362 | -0.59 | -4.84, 3.66 | 0.786 | 42 | 0.15 | -0.32, 0.62 | 0.526 |

| | | |
|---|---|---|
| Observations | 955 | 8140 |
| $R^2$ / $R^2$ adjusted | 0.162 / 0.147 | 0.101 / 0.099 |

Model adjusted by age, ethnicity, sex, taking inhibitors of CYP2D6, taking antidiabetics and BMI, Normal metabolisers of CYP2D6: tricyclics diabetes 703, amitriptyline diabetes = 638, Normal metabolisers of CYP2C19: tricyclics diabetes = 345, amitriptyline = 322

**Supplementary table 21. Antipsychotics regression model. Association between CYP2D6 metabolic phenotype and HbA1c – additional detail**

| | Antipsychotics | | |
|---|---|---|---|
| *Predictors* | *Estimates* | *CI* | *p* |
| CYP2D6 IM | -0.02 | -0.58,0.53 | 0.930 |
| CYP2D6 PM | -0.93 | -2.01,0.16 | 0.093 |
| Takes CYP2D6 inhibitor | 0.59 | -0.43,1.61 | 0.260 |
| Sex: Male | 0.40 | -0.07,0.88 | 0.097 |
| Age at recruitment | 0.09 | 0.06,0.12 | **<0.001** |
| Ethnicity: Admix Caucasian | 0.78 | -0.67,2.23 | 0.291 |
| Ethnicity: African | 3.81 | 2.49,5.13 | **<0.001** |
| Ethnicity: East Asian | 2.31 | -1.21,5.83 | 0.198 |
| Ethnicity: Other | 0.94 | -0.70,2.58 | 0.263 |
| Ethnicity: South Asian | 3.72 | 2.19,5.26 | **<0.001** |
| Diabetes | 4.55 | 3.13,5.97 | **<0.001** |
| BMI | 0.19 | 0.15,0.23 | **<0.001** |
| Antidiabetics | 14.18 | 12.55,15.81 | **<0.001** |
| Observations | 2699 | | |
| $R^2$ / $R^2$ adjusted | 0.449 / 0.446 | | |
